# Geographic prioritization of child nutrition services in Bangladesh: comparing stunting and wasting indicators using the 2025 Multiple Indicator Cluster Survey

**DOI:** 10.64898/2026.10.02.26364425

**Authors:** S. M. Ashraf Ul Hoque, Md Khairul Islam, Mohammad Khalid Hasan, Mohiuddin Sharif, Tanzin Naher, Bilkis Banu, Zenat Zebin Hossain, Kamran ul Baset

## Abstract

**Background:** Bangladesh’s progress on stunting leaves policymakers with a practical question about wasting: where should they focus limited resources? The country met its 2025 stunting target, yet child wasting rose from 9.8% in 2019 to 12.9% in 2025. We assessed whether prioritizing districts with the highest stunting prevalence would include a greater share of children with wasting than proportional allocation at the same child-population coverage.

**Methods:** We used the 2025 Bangladesh Multiple Indicator Cluster Survey to rank all 64 districts by stunting or wasting prevalence. Among 22,311 children under five with valid weight-for-height measurements, we estimated the share of children with wasting who lived in the highest-ranked districts. We compared rankings with population-proportional allocation at 10%, 20%, and 30% child-population coverage, interpolating within the last district to match coverage exactly. We estimated uncertainty using 2,000 paired survey bootstrap replicates and assessed how well district wasting rankings from 2019 carried forward to 2025.

**Results:** At 20% coverage, districts ranked highest for stunting contained 19.5% of children with wasting (95% CI 17.8 to 22.6), a difference of −0.5 percentage points from proportional allocation (95% CI −2.2 to 2.6). Differences were also small at 10% (−0.3 points, 95% CI −1.6 to 1.2) and 30% coverage (1.3 points, 95% CI −2.3 to 3.3). The four stunting hotspot districts contained 5.9% of all children and 5.9% of children with wasting. At 20% coverage, ranking districts by 2025 wasting prevalence captured 26.3% of children with wasting in-sample and 24.3% after correction for optimism. The 2019 wasting ranking captured 22.9% (95% CI 19.0 to 24.2).

**Conclusions:** A stunting map should not guide where Bangladesh prioritizes districts for child wasting. High-stunting districts held wasted children only in proportion to their population. Wasting rankings from 2019 kept an uncertain advantage. Wasting prioritization needs its own, more frequent data.

## Introduction

Child stunting and wasting both raise the risk of death [1,2], and children with both deficits face the highest risk [3]. Reducing both is part of Sustainable Development Goal target 2.2. The global nutrition targets for 2030 call for wasting below 5% [4]. In Bangladesh, the 2025 Multiple Indicator Cluster Survey (MICS) found 24.0% of children under five stunted and 12.9% wasted, up from 9.8% wasted in 2019 [5]. The Second National Plan of Action for Nutrition set 2025 targets of 25% for stunting and less than 8% for wasting [6]. The stunting target was met, but the wasting target was not [5,6].

National nutrition planners and district health managers must decide where to add effort when resources are limited. One common input is a map of undernutrition by area. The official undernutrition maps of Bangladesh, produced by the Bangladesh Bureau of Statistics and the World Food Programme, estimated stunting and underweight for every district and upazila [7]. They state that such maps were requested to aid policymaking and program targeting [7]. They did not map wasting because it varies by season [7]. In 2025, an Integrated Food Security Phase Classification (IPC) acute malnutrition analysis classified 18 districts and the Rohingya refugee populations by phase of severity [8]. It did not rank all 64 districts by wasting prevalence. Large programs have also been placed in high-stunting areas. Suchana, a European Union co-funded program, worked in Sylhet and Moulvibazar districts with the aim of reducing stunting in children under two [9]. Its published factsheet does not state why these two districts were chosen [9]. The government’s wasting commitment for 2021 to 2025 names national guidelines for community-based management of acute malnutrition among its instruments [10]. In the documents we consulted, we found no statement that wasting services are placed according to stunting maps. Using a stunting map to place effort against wasting is therefore a plausible proposed strategy, not a documented practice.

There are reasons to doubt that such a strategy would work. Stunting reflects long-term growth failure, while wasting reflects recent weight loss and can change within months [11]. Subnational maps across low- and middle-income countries show large differences within countries for both conditions [12]. Studies in Bangladesh have mapped each condition at the district or a finer level [13,14]. Concurrent wasting and stunting is common and dangerous [15,16]. Even so, only 31.2% of wasted children in Bangladesh in 2025 were also stunted (902 of 2,881 with both measurements; Table 1). None of these studies asked the planning question directly. That question is how much of the wasting burden a stunting-based district list would include, at a given share of the child population.

**Table 1.**
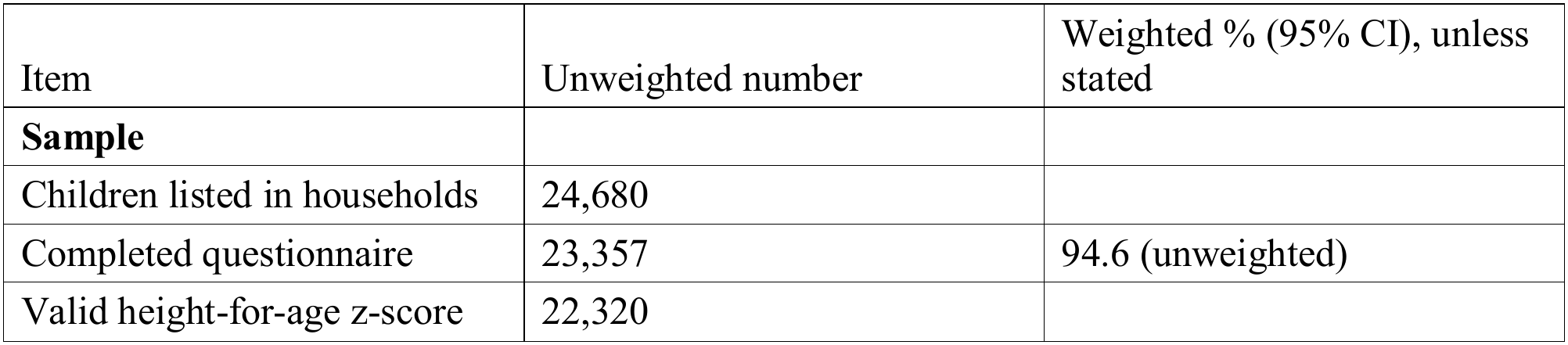
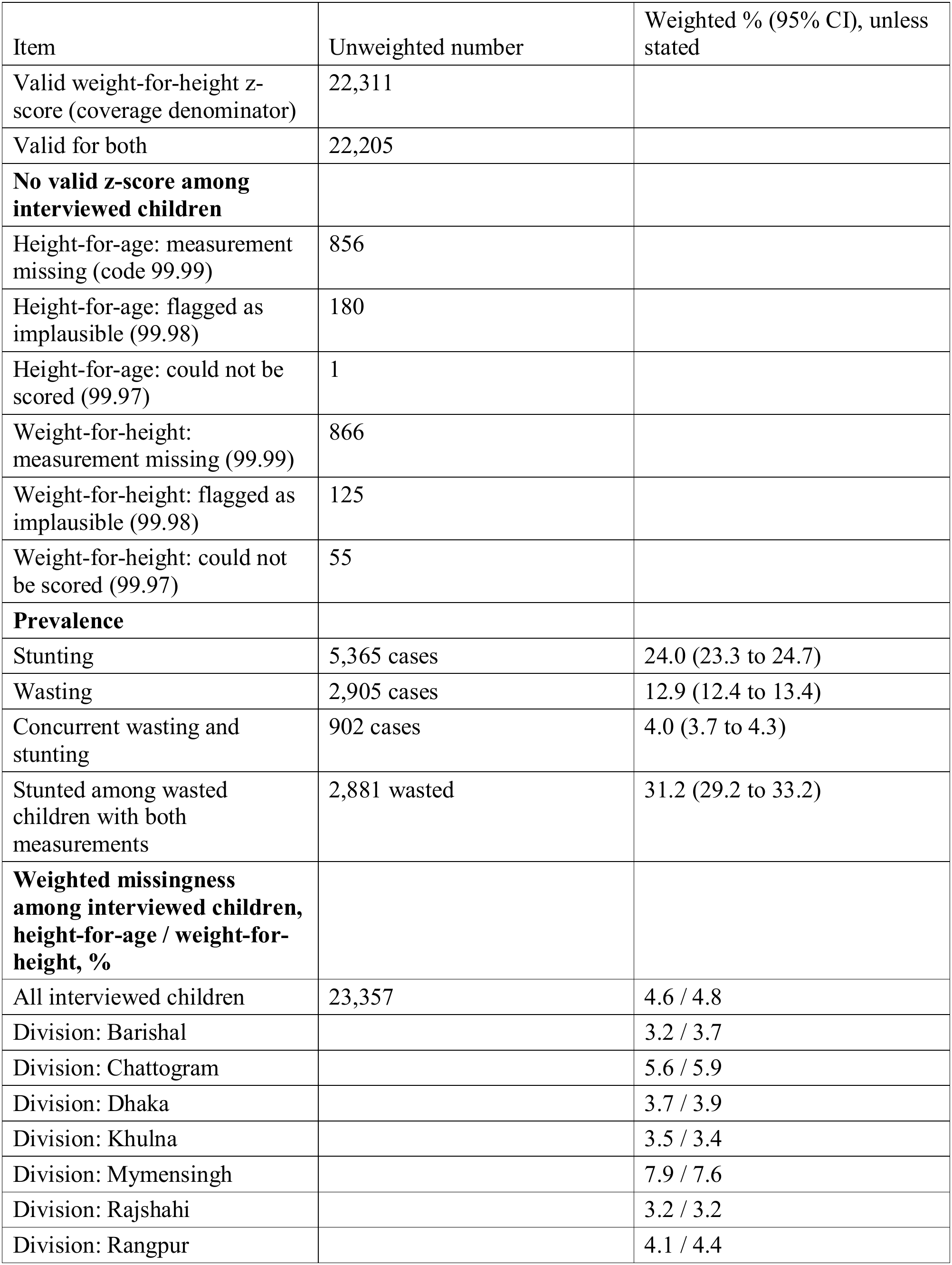
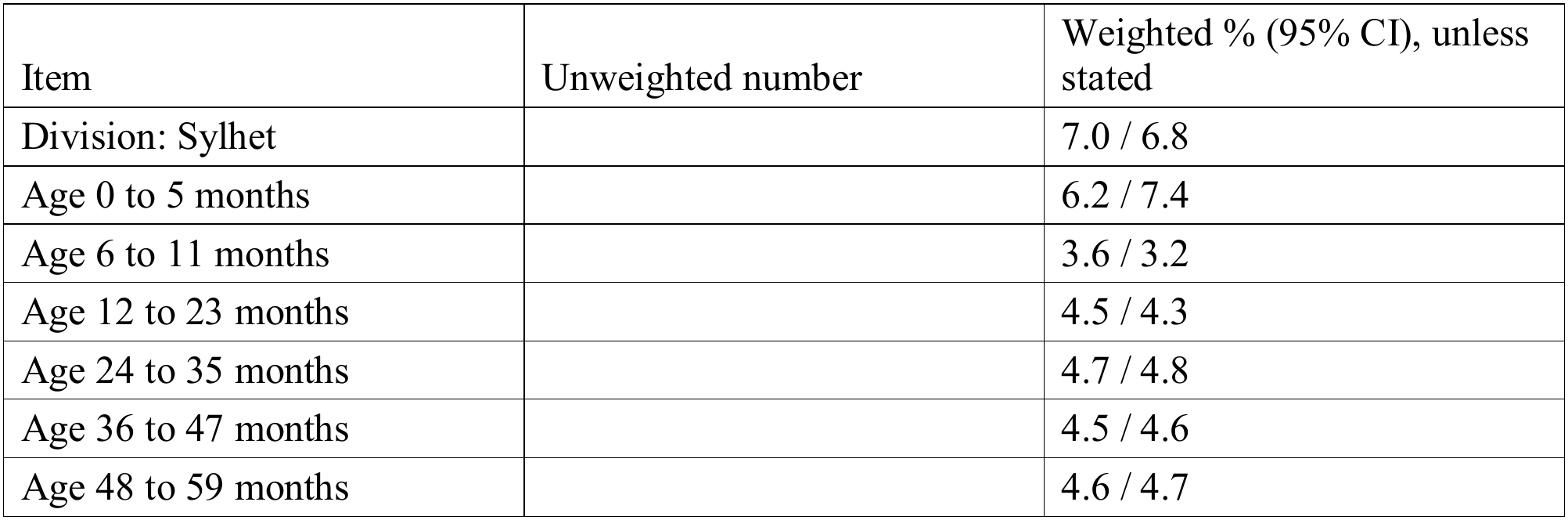
Analytic population, prevalence and missing anthropometry, Bangladesh MICS 2025.

| Item | Unweighted number | Weighted % (95% CI), unless stated |
|---|---|---|
| **Sample** |  |  |
| Children listed in households | 24,680 |  |
| Completed questionnaire | 23,357 | 94.6 (unweighted) |
| Valid height-for-age z-score | 22,320 |  |
| Valid weight-for-height z-score (coverage denominator) | 22,311 |  |
| Valid for both | 22,205 |  |
| **No valid z-score among interviewed children** |  |  |
| Height-for-age: measurement missing (code 99.99) | 856 |  |
| Height-for-age: flagged as implausible (99.98) | 180 |  |
| Height-for-age: could not be scored (99.97) | 1 |  |
| Weight-for-height: measurement missing (99.99) | 866 |  |
| Weight-for-height: flagged as implausible (99.98) | 125 |  |
| Weight-for-height: could not be scored (99.97) | 55 |  |
| **Prevalence** |  |  |
| Stunting | 5,365 cases | 24.0 (23.3 to 24.7) |
| Wasting | 2,905 cases | 12.9 (12.4 to 13.4) |
| Concurrent wasting and stunting | 902 cases | 4.0 (3.7 to 4.3) |
| Stunted among wasted children with both measurements | 2,881 wasted | 31.2 (29.2 to 33.2) |
| **Weighted missingness among interviewed children, height-for-age / weight-for-height, %** |  |  |
| All interviewed children | 23,357 | 4.6 / 4.8 |
| Division: Barishal |  | 3.2 / 3.7 |
| Division: Chattogram |  | 5.6 / 5.9 |
| Division: Dhaka |  | 3.7 / 3.9 |
| Division: Khulna |  | 3.5 / 3.4 |
| Division: Mymensingh |  | 7.9 / 7.6 |
| Division: Rajshahi |  | 3.2 / 3.2 |
| Division: Rangpur |  | 4.1 / 4.4 |
| Division: Sylhet |  | 7.0 / 6.8 |
| Age 0 to 5 months |  | 6.2 / 7.4 |
| Age 6 to 11 months |  | 3.6 / 3.2 |
| Age 12 to 23 months |  | 4.5 / 4.3 |
| Age 24 to 35 months |  | 4.7 / 4.8 |
| Age 36 to 47 months |  | 4.5 / 4.6 |
| Age 48 to 59 months |  | 4.6 / 4.7 |

We therefore compared rules for choosing districts at the same child-population coverage. Our primary comparison was between a stunting-based list, a wasting-based list and population-proportional allocation. “Included” here means only that wasted children live in the selected districts. It does not mean that they are screened, treated or helped. A list built from one survey also depends on sampling error, so we asked how stable district lists are under survey uncertainty. Finally, we asked whether wasting rankings from the 2019 survey still selected high-wasting districts in 2025. Spatial clustering analyses, which identified the stunting hotspot districts, are reported as secondary.

## Methods

### Study design and data

This is a cross-sectional analysis of the Bangladesh MICS 2025, conducted by the Bangladesh Bureau of Statistics with UNICEF Bangladesh [5]. The survey used a stratified two-stage cluster sample of enumeration areas and households. Fieldwork ran from 24 February to 30 June 2025, and the household response rate was 99.6% [5]. For the temporal assessment, we used the Bangladesh MICS 2019, which had a similar design and the same anthropometric indicators [17].

### Population and outcomes

The population was all children under five listed in the 2025 survey, so no sample size calculation was needed. Wasting was defined as a weight-for-height z-score below −2, and stunting as a height-for-age z-score below −2 (2006 World Health Organization child growth standards). We used the z-scores released by MICS (variables HAZ2 and WHZ2), so wasting was not defined by mid-upper arm circumference. A z-score was valid if MICS had calculated it and had not assigned an exclusion code (S1 Appendix). Children with a valid wasting measurement formed the denominator for all coverage measures. The 2019 data were prepared with the same rules.

### Districts

We merged the Dhaka North, Dhaka South and Chattogram city corporations into their parent districts, giving 64 districts in both surveys. District names were matched to geoBoundaries [18].

### Selection rules

We compared four rules for choosing districts. Rule A, the strategy of interest, ranked districts by 2025 stunting prevalence and added them in rank order. Rule B ranked districts by 2025 wasting prevalence in the same way. Because it is judged on the data used to rank, it is an in-sample benchmark expected to look better than it would in practice. Rule C was population-proportional allocation, which selects the same fraction of children in every district. Rule D used the four districts meeting the nominal Getis-Ord Gi* hotspot threshold (z > 1.96): Moulvibazar, Netrokona, Sunamganj and Sylhet. Three also met the 5% false discovery rate criterion. Secondary rules ranked districts by the number of stunted children and by 2019 stunting prevalence.

### Coverage and burden measures

Coverage is the weighted share of children living in the selected districts, and burden capture is the corresponding share of wasted children. Enrichment, burden capture divided by coverage, equals 1 when a list includes wasted children exactly in proportion to its share of all children, as under rule C. We also report burden capture minus coverage in percentage points. We used coverage scenarios of about 10%, 20% and 30% of children as reference points, not recommended program sizes. Districts were added until coverage first reached each target. For comparison at exactly the same coverage, we interpolated within the last district, assuming its average wasting prevalence (coverage-matched results).

### Survey uncertainty

We estimated uncertainty with a stratified rescaling bootstrap for complex surveys [19], using 2,000 replicates. Each replicate repeated district prevalence, ranking, selection, coverage and burden. All rules used the same replicates, so differences between rules are paired. We report percentile 95% confidence intervals (CIs), which include the chance that a different set of districts would have been chosen. We also report the share of replicates in which each district was selected (S1 Table; S1 Fig). For rule B, we omit percentile intervals for burden capture and enrichment under repeated selection, because ranking by observed prevalence maximizes the observed share by construction. For the stunting ranking, we also built a simultaneous 95% band over coverage. Fixed-list intervals, a check with all household primary sampling units (PSUs) and further details are in S1 Appendix. We made no adjustment for multiple comparisons, and contrasts other than the primary comparison are exploratory.

### Selection optimism

Ranking and judging districts on 2025 wasting also captures sampling noise. Our main check of this optimism was a temporal assessment. We ranked districts by wasting prevalence in MICS 2019 and judged the resulting lists against 2025 wasting, resampling the 2019 sample independently. This mirrors a planner using the most recent survey. As a supporting check, we estimated and subtracted bootstrap optimism [20] (S1 Appendix).

### Sensitivity analyses

We described missing anthropometry by division, age group, wealth quintile and area with design-based tests. We then repeated the selection analyses with two adjustments, both recomputed in every replicate (S1 Appendix). First, because fieldwork reached districts in different months, we standardized district wasting for month of measurement. Second, we reweighted children with a valid z-score for missing anthropometry within cells of district by age group. We also ran a reverse analysis: the share of stunted children included by wasting-based selection.

### Secondary spatial and environmental analyses

We tested clustering of district prevalence with Moran’s I [21] and identified hotspots with the Getis-Ord Gi* statistic [22]. False discovery rate correction used the Benjamini-Hochberg method. Survey-weighted logistic models related six cluster-level environmental and access factors to each outcome (S1 Appendix). The environmental models played no role in district selection (S2 and S3 Tables; S2 to S4 Figs).

### Prespecification

The selection rules and the analyses built on them were not prespecified. We developed them after an initial spatial analysis of the same data, which had reported lists at 10%, 20% and 30% coverage built from stunting rankings. Compared with that analysis, this paper adds equal-coverage comparisons with a proportional benchmark, uncertainty in which districts are selected, an optimism correction and a temporal assessment.

### Software and reporting

The selection analyses were run in Stata version 17. We used R version 4.6 for the figures, the secondary analyses and the check of missing codes. Reporting follows the Strengthening the Reporting of Observational Studies in Epidemiology guideline for cross-sectional studies (S1 Checklist).

### Ethics statement

This secondary analysis used publicly available survey data. We accessed the data on 24 September 2026 (MICS 2019), 26 September 2026 (MICS 2025) and 28 September 2026 (cluster locations). The files were anonymized, and the authors could not identify participants during or after data collection. For MICS 2025, the icddr,b Ethical Review Committee approved the protocol (ref. PR-24113), and the implementing agencies obtained informed consent. For MICS 2019, a government technical committee led by the Bangladesh Bureau of Statistics approved the protocol, and respondents gave verbal consent [17]. No separate ethical review was needed. MICS displaced cluster locations, used only in the secondary analyses, before release [23].

## Results

### Sample and missing data

Of 24,680 listed children, 23,357 (94.6%) had a completed questionnaire (Table 1). After children without a valid z-score were excluded, 22,320 remained for stunting and 22,311 for wasting, with 22,205 measured for both. In the text, counts are unweighted and percentages are survey-weighted. The analysis covered 64 districts, 131 strata and 3,146 PSUs.

Children without a completed questionnaire have zero weight in the released file because MICS weights adjust for questionnaire nonresponse. Counts are unweighted; percentages are weighted unless stated. Missing codes are as assigned by MICS, and recomputation with the WHO anthro R package confirmed every checkable 99.98 flag (S1 Appendix).

Weighted missingness of the height-for-age z-score was 4.6% (1,037 of 23,357 interviewed children). It was highest in the Mymensingh and Sylhet divisions and in infants under 6 months, and it did not vary by wealth quintile or residence (Table 1; S1 Appendix).

Weighted prevalence was 24.0% for stunting (5,365 of 22,320; 95% CI 23.3 to 24.7) and 12.9% for wasting (2,905 of 22,311; 95% CI 12.4 to 13.4). Concurrent wasting and stunting affected 4.0% (902 of 22,205; 95% CI 3.7 to 4.3). Among wasted children with both measurements, 31.2% (902 of 2,881; 95% CI 29.2 to 33.2) were also stunted (Table 1).

### District distributions

District stunting prevalence ranged from 17.0% in Kushtia (51 of 306) to 37.3% in Sylhet (145 of 389). District wasting prevalence ranged from 6.3% in Manikganj (18 of 268) to 20.0% in Naogaon (40 of 211) (Fig 1). The two showed no linear association (Pearson r −0.01 and Spearman 0.06 across 64 districts, given descriptively). Stunting was highest in the northeast and south, and wasting was highest mostly in the northwest and west (Fig 1).

**Fig 1.**
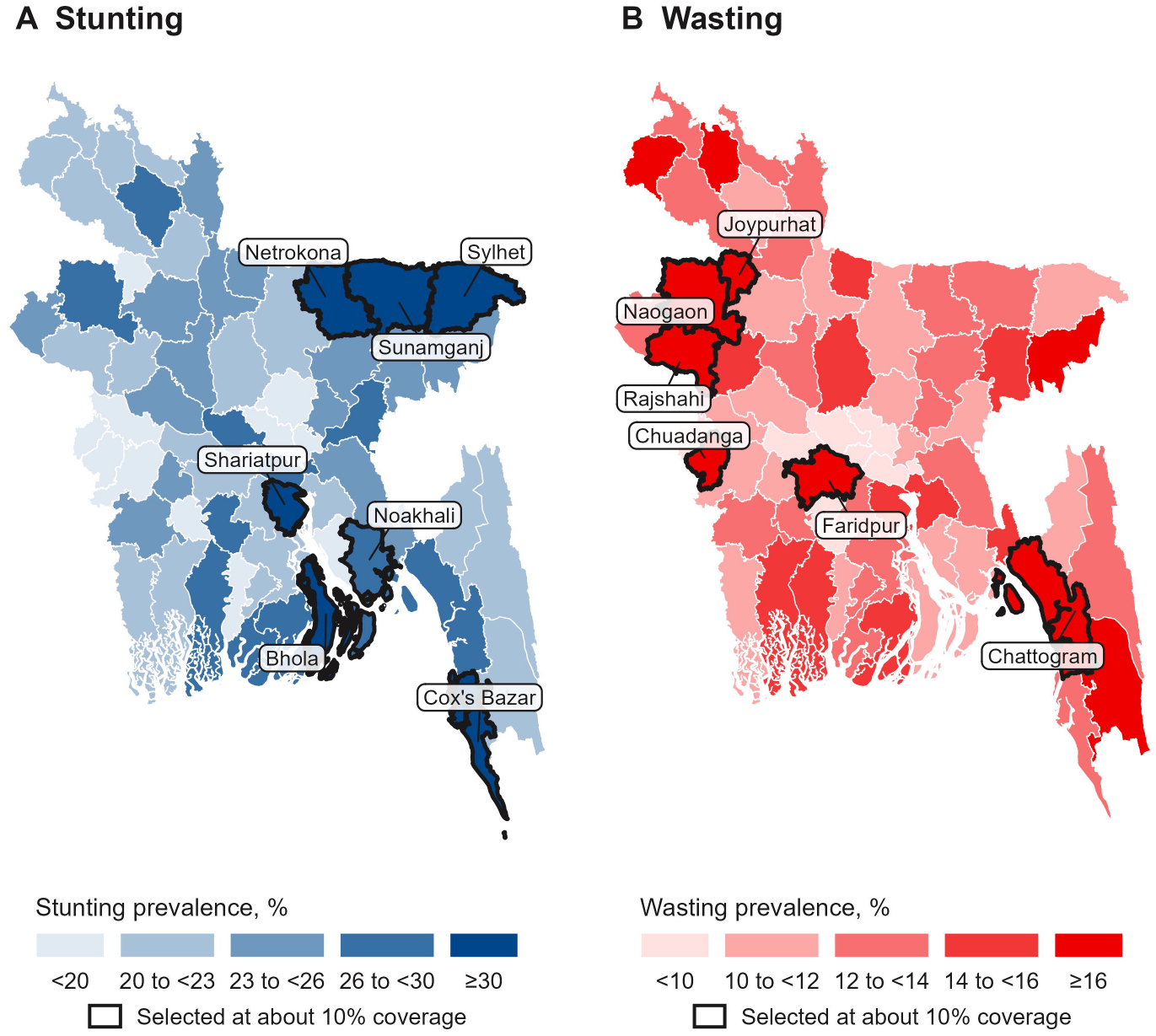
District prevalence of stunting (A) and wasting (B), Bangladesh MICS 2025. Each map shows 64 districts, with city corporations merged into their parent districts. Outlined districts were selected by ranking on that indicator until about 10% of children were covered. Prevalence is survey-weighted. District 95% confidence intervals are in S1 Table, panel F. District boundaries are from geoBoundaries [18].

District wasting estimates were imprecise, with a median 95% CI width of 7.5 percentage points (range 4.0 to 12.0; S1 Table, panel F). Between 2019 and 2025, district wasting prevalence correlated weakly (r 0.32), and district stunting prevalence correlated moderately (r 0.45).

### Primary comparison

At the three coverage levels, districts ranked by stunting included wasted children at about their population share (Table 2, Fig 2). For example, the first 13 districts by stunting held 20.1% of children (4,726 of 22,311) and 19.6% of wasted children (595 of 2,905). Their enrichment was 0.98 (95% CI 0.89 to 1.13).

**Fig 2.**
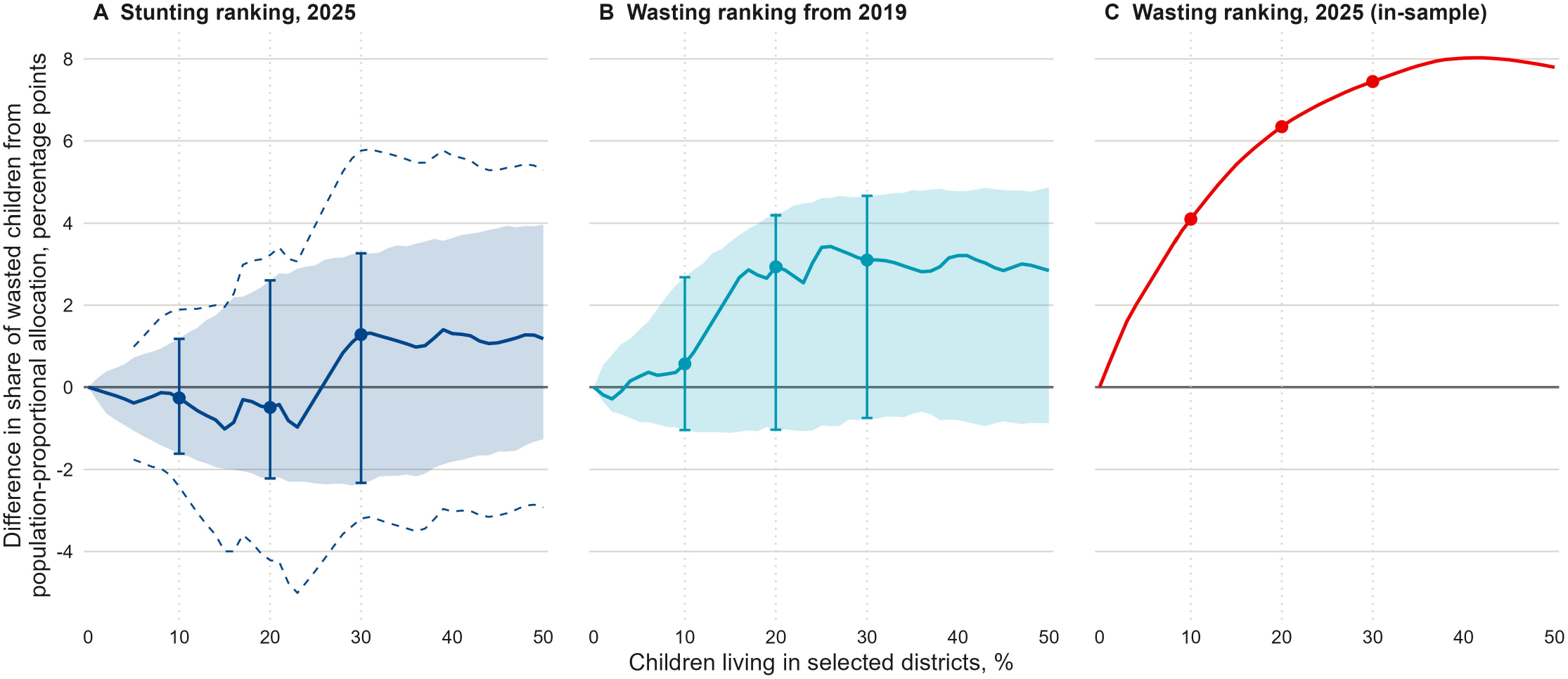
Difference in the share of wasted children from population-proportional allocation, by ranking rule, Bangladesh MICS 2025. Each panel adds districts in rank order up to 50% child-population coverage. Values above zero mean the selected districts held more wasted children than their share of all children. Shaded bands are pointwise 95% bootstrap intervals from 2,000 paired replicates that repeat estimation, ranking and selection. Dashed lines in panel A are a simultaneous 95% band over 5% to 95% coverage. Points and bars mark 10%, 20% and 30% coverage. The 2025 wasting ranking (C) is an in-sample benchmark without intervals, because its replicate intervals are biased upward.

**Table 2.**
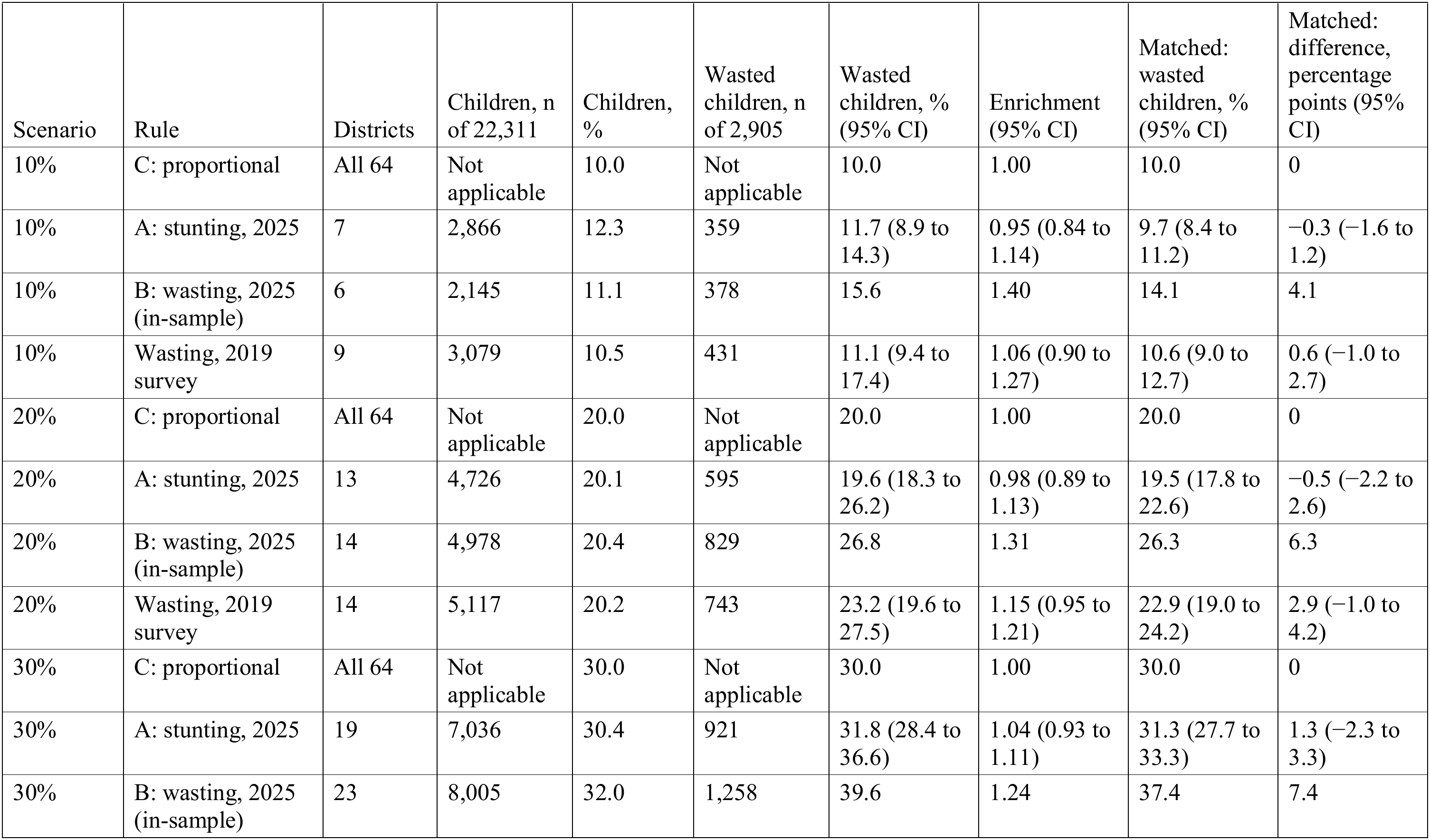
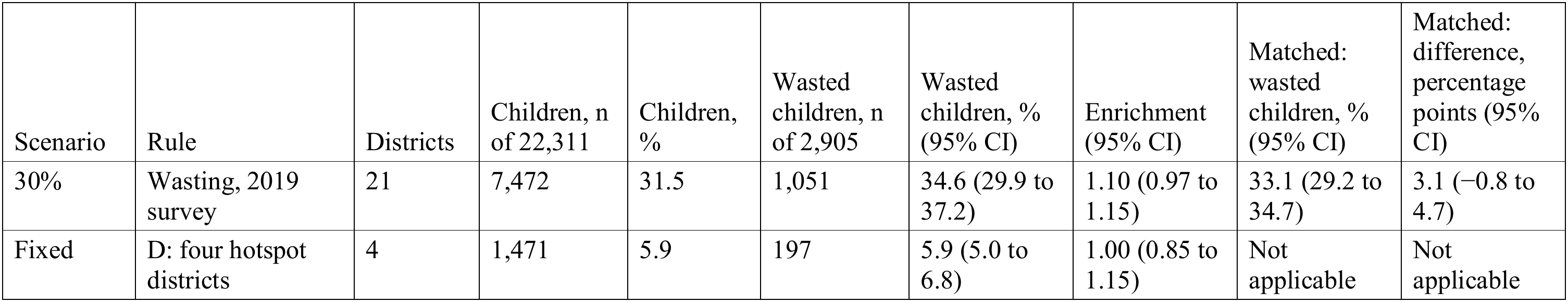
Wasted children living in districts selected by each rule, at about 10%, 20% and 30% coverage of children, Bangladesh MICS 2025.

| Scenario | Rule | Districts | Children, n of 22,311 | Children, % | Wasted children, n of 2,905 | Wasted children, % (95% CI) | Enrichment (95% CI) | Matched: wasted children, % (95% CI) | Matched: difference, percentage points (95% CI) |
|---|---|---|---|---|---|---|---|---|---|
| 10% | C: proportional | All 64 | Not applicable | 10.0 | Not applicable | 10.0 | 1.00 | 10.0 | 0 |
| 10% | A: stunting, 2025 | 7 | 2,866 | 12.3 | 359 | 11.7 (8.9 to 14.3) | 0.95 (0.84 to 1.14) | 9.7 (8.4 to 11.2) | −0.3 (−1.6 to 1.2) |
| 10% | B: wasting, 2025 (in-sample) | 6 | 2,145 | 11.1 | 378 | 15.6 | 1.40 | 14.1 | 4.1 |
| 10% | Wasting, 2019 survey | 9 | 3,079 | 10.5 | 431 | 11.1 (9.4 to 17.4) | 1.06 (0.90 to 1.27) | 10.6 (9.0 to 12.7) | 0.6 (−1.0 to 2.7) |
| 20% | C: proportional | All 64 | Not applicable | 20.0 | Not applicable | 20.0 | 1.00 | 20.0 | 0 |
| 20% | A: stunting, 2025 | 13 | 4,726 | 20.1 | 595 | 19.6 (18.3 to 26.2) | 0.98 (0.89 to 1.13) | 19.5 (17.8 to 22.6) | −0.5 (−2.2 to 2.6) |
| 20% | B: wasting, 2025 (in-sample) | 14 | 4,978 | 20.4 | 829 | 26.8 | 1.31 | 26.3 | 6.3 |
| 20% | Wasting, 2019 survey | 14 | 5,117 | 20.2 | 743 | 23.2 (19.6 to 27.5) | 1.15 (0.95 to 1.21) | 22.9 (19.0 to 24.2) | 2.9 (−1.0 to 4.2) |
| 30% | C: proportional | All 64 | Not applicable | 30.0 | Not applicable | 30.0 | 1.00 | 30.0 | 0 |
| 30% | A: stunting, 2025 | 19 | 7,036 | 30.4 | 921 | 31.8 (28.4 to 36.6) | 1.04 (0.93 to 1.11) | 31.3 (27.7 to 33.3) | 1.3 (−2.3 to 3.3) |
| 30% | B: wasting, 2025 (in-sample) | 23 | 8,005 | 32.0 | 1,258 | 39.6 | 1.24 | 37.4 | 7.4 |
| 30% | Wasting, 2019 survey | 21 | 7,472 | 31.5 | 1,051 | 34.6 (29.9 to 37.2) | 1.10 (0.97 to 1.15) | 33.1 (29.2 to 34.7) | 3.1 (−0.8 to 4.7) |
| Fixed | D: four hotspot districts | 4 | 1,471 | 5.9 | 197 | 5.9 (5.0 to 6.8) | 1.00 (0.85 to 1.15) | Not applicable | Not applicable |

Scenarios correspond to about 10%, 20% and 30% of children. The n columns give unweighted counts in the selected districts. Their denominators l children with a valid weight-for-height z-score (22,311) and all wasted children (2,905). Percentages are survey-weighted, so they can differ n divided by its denominator. Rule C selects a fraction of children in every district and has no district counts. Districts, Children and Wasted en refer to whole-district lists, which add districts in rank order until coverage first reaches the scenario. Matched columns give coverage-matched estimates at exactly the scenario coverage; the difference is from rule C in percentage points. Coverage-matched columns interpolate within st district, assuming its average wasting prevalence. Intervals are percentile intervals from 2,000 paired bootstrap replicates that repeat estimation, ranking and selection. The 2019 ranking was resampled independently of the 2025 data. Enrichment is the share of wasted children divided by the share of all children in the selected districts. Rule B is an in-sample benchmark. Ranking by observed wasting maximizes the observed by construction, so its percentile intervals lack nominal coverage and are not shown. Its optimism-corrected coverage-matched shares were 12.7%, 24.3% and 34.7% (S1 Table, panel D). Rule C includes wasted children at exactly the coverage fraction by construction; random selection of en would give this share on average. Rule D comprises the four nominal Getis-Ord Gi* stunting hotspots from our analysis of the same data lvibazar, Netrokona, Sunamganj and Sylhet). The 2019 rankings were applied to 2025 child-population shares.

Coverage-matched estimates gave the same picture (Table 2). At exactly 10%, 20% and 30% coverage, stunting-ranked districts included 9.7%, 19.5% and 31.3% of wasted children. These differed from population-proportional allocation by −0.3 (95% CI −1.6 to 1.2), −0.5 (−2.2 to 2.6) and 1.3 (−2.3 to 3.3) percentage points. At these three levels, the intervals excluded gains or losses larger than about 3.3 percentage points. Across the whole curve, the simultaneous band was wider: −4.2 to 3.2 points at 20% coverage and −3.2 to 5.8 at 30% (Fig 2A).

Districts ranked by 2025 wasting included 14.1%, 26.3% and 37.4% of wasted children at the same three coverage levels. Ranking by observed wasting maximizes this share by construction, so these in-sample figures have no intervals and overstate achievable performance.

The four stunting hotspot districts held 5.9% of children (1,471 of 22,311) and 5.9% of wasted children (197 of 2,905). Their enrichment was 1.00 (95% CI 0.85 to 1.15; Table 2).

### Uncertainty and stability of district lists

District lists changed between replicates. Sylhet and Sunamganj entered the 10% stunting list in 99% or more of the 2,000 replicates. No district entered the 10% wasting list in more than 83% (S1 Table, panel G; S1 Fig). Stunting-ranked districts included fewer wasted children than population-proportional allocation in 65%, 55% and 39% of replicates at 10%, 20% and 30% coverage (S1 Table, panel A).

### Selection optimism and the temporal assessment

At matched coverage, 2019 wasting rankings judged against 2025 data included 10.6% (95% CI 9.0 to 12.7), 22.9% (19.0 to 24.2) and 33.1% (29.2 to 34.7) of wasted children. Their enrichments were lower than the in-sample 2025 values, and every interval included 1 (S1 Table, panel A). When both rankings came from 2019, the exploratory paired differences favored the wasting ranking, but the intervals included zero at 10% and 20% coverage (S1 Table, panel B).

Bootstrap optimism of the in-sample wasting ranking was 1.4, 2.1 and 2.7 percentage points (S1 Table, panel D). At 20% coverage, the wasting ranking therefore included 26.3% of wasted children in-sample, 24.3% after optimism correction and 22.9% when ranked with 2019 data. Estimated optimism for the stunting ranking was 0.15 percentage points or less. For the in-sample wasting ranking at 30% coverage, the bootstrap percentile range (37.5% to 40.6%) lay above the observed share (37.4%).

### Sensitivity analyses

Month standardization and response reweighting changed the results little. Coverage-matched enrichment of the stunting ranking was 0.99, 0.98 and 1.04 after standardization and 0.98, 0.97 and 1.04 after reweighting (S1 Table, panel A). Ranking by the number of stunted children gave similar results. In the reverse analysis, districts ranked by wasting included stunted children in proportion to their population share (S1 Table, panel A).

### Secondary spatial and environmental results

Stunting clustered across districts (Moran’s I 0.21, P=0.003), with four nominal hotspot districts in the northeast, three of which passed false discovery rate correction (S2 Fig). District clustering of wasting was not detected (I 0.05, P=0.17). After adjustment for six environmental and access factors, stunting clustering was no longer detected. The role of these factors could not be separated from region (S2 Table, panels B and C).

## Discussion

In Bangladesh in 2025, districts chosen by stunting prevalence included wasted children at about their share of the child population. This held at 10%, 20% and 30% coverage, for the four-district stunting hotspot list, and after month standardization and response reweighting. At each of these three levels, the intervals were compatible with differences of up to about 3 percentage points either way. Across all coverage levels up to 50%, the simultaneous band allowed differences of up to about 6 points. A stunting-based list therefore performed about as well as population-proportional allocation for wasting, with no clear advantage. Wasting-based rankings did better within the 2025 survey. Much of that advantage was lost when 2019 rankings were applied to 2025 data. The data cannot separate how much of this loss came from selection optimism, change over six years or different fieldwork months. In 2019, child interviews ran from January to early June, against February to June in 2025. The remaining gain was uncertain.

### Comparison with other studies

In our data, district stunting and wasting prevalence showed no linear association. A correlation across all districts cannot show how many wasted children the highest-ranked districts hold, and the coverage curves show this directly. Our finding agrees with work showing that stunting and wasting have different geographies. Across low- and middle-income countries, subnational maps have shown large and separate within-country patterns for each condition [12]. Analyses of cohort data show that wasting often comes before stunting and varies by season [16]. Reviews have argued that research and programs should treat the two conditions as related but distinct [24]. Studies in Bangladesh have produced district-level or finer maps of each condition [13,14,25]. A study in Ethiopia found that local clusters of stunting and wasting did not coincide [26]. In Nepal, a bivariate model of 2016 survey data found higher stunting risk in Lumbini and Karnali provinces, and higher wasting risk in Madhesh and Province 1 [27]. Across 640 districts of India in 2015-16, both stunting and wasting clustered spatially [28]. Neither study asked how many wasted children a stunting-based list would include. We add a planning measure: the share of wasted children included at a given coverage, with an explicit check of selection optimism. The official undernutrition maps left out wasting because of seasonality [7]. Our results support that caution. In this survey, a stunting ranking did not substitute for wasting data.

### Meaning for planners

The finding answers a narrow question. Suppose national or district planners use a stunting map to decide where to add attention to wasting. In this survey, a stunting ranking added little information about wasting beyond population size. A stunting map remains a guide for stunting work, although our evidence for this is partly in-sample. Stunting-ranked districts held more stunted children than their population share in the same survey that ranked them. District stunting prevalence also correlated moderately between 2019 and 2025.

A wasting map from a single survey is not a ready alternative. District wasting estimates rested on a few hundred children each, and their intervals were wide. Lists built from them changed from replicate to replicate. Rankings from 2019 kept only part of their value six years later, and district wasting correlated weakly between the two surveys. A planner choosing between indicators at one time faces a different comparison, with both rankings from 2019. In that exploratory comparison, the 2019 wasting ranking included more 2025 wasted children than the 2019 stunting ranking. The differences were 1.5 (95% CI −1.1 to 3.7), 3.9 (−0.4 to 6.5) and 6.0 (0.5 to 8.0) percentage points at 10%, 20% and 30% coverage. Against proportional allocation, the advantage of the 2019 wasting ranking remained uncertain at all three levels. Wasting can also change within months [11], and MICS measured each district during only part of a single February-to-June season. For wasting, district rankings from one household survey are better treated as a signal for further assessment than as a basis for allocation.

These results do not support withdrawing effort from high-stunting districts. Stunting-ranked districts such as Sylhet and Sunamganj had wasting prevalence close to the national average, so they held a population-proportional share of wasted children. The choice between rules is also an equity question. A wasting ranking would move attention toward western districts and away from the northeast, where stunting is highest. A combined approach could keep stunting-based effort and add a wasting signal from a source measured more often. Such a source, for example routine screening records or repeated surveys, needs evaluation of data quality and of the services that follow.

The study has limited scope. It measured geographic inclusion: whether wasted children live in a selected district. It did not measure surveillance participation, access to services, screening, treatment coverage, recovery or deaths averted. A district list that includes many wasted children helps them only if services reach them. Nor do our results say how much any district should receive. Costs, existing service coverage and case severity all bear on that choice. Our estimates come from a household survey that measures prevalence, not incidence. A child may be wasted more than once in a year, so prevalence-based shares are not annual caseloads.

### Strengths and limitations

The study used a large, recent and nationally representative survey with standard anthropometry. The bootstrap repeated estimation, ranking and selection in each replicate, so intervals reflect uncertainty in which districts are chosen. All rules were compared on the same replicates. We addressed selection optimism directly with an independent earlier survey.

There are several limitations. First, none of the selection analyses, including the temporal check, was prespecified. Second, the analysis was conducted at the district level. Programs often work at the upazila or union level, where the overlap of stunting and wasting may differ. MICS cannot estimate prevalence reliably at those levels. Third, the temporal assessment spans six years, a pandemic and a rise in national wasting. It may understate how well a recent wasting survey would perform, and it says nothing about routine data. The 2019 orderings were also applied to the 2025 child population, which a planner in 2019 could not have done exactly. Fourth, month standardization assumed the same seasonal pattern in every district and covered only February to June. Districts were measured in different months, and fieldwork timing correlated with district wasting (S2 Table, panel D). National monthly prevalence therefore partly reflects which districts were measured each month, so standardization may not separate season from place. Fifth, some interviewed children lacked a valid z-score, more often in the Sylhet and Mymensingh divisions and among infants. Reweighting assumed that missing children resembled measured children of the same age in the same district. Sixth, wasting was defined by weight-for-height only. Programs often identify wasting by mid-upper arm circumference [11], which detects a partly different group of children. Seventh, interpolated estimates assume that part of a district can be included at the district’s average prevalence. Whole-district results, which need no such assumption, gave the same conclusions.

## Conclusions

A stunting map should not be used as a proxy for deciding where Bangladesh focuses effort against child wasting. At 10%, 20% and 30% child-population coverage, districts ranked by stunting held wasted children only in proportion to their share of all children, as population-proportional allocation would. Wasting rankings did better within one survey, but much of that advantage was lost when 2019 rankings were applied to 2025. District prioritization for wasting therefore needs its own data, measured more often than a household survey every few years. Routine screening records could serve once their quality is evaluated. Stunting-based effort should continue for stunting itself. These findings describe where wasted children lived, not whether services reached them.

## Data Availability

The 2025 Bangladesh Multiple Indicator Cluster Survey (MICS) data, including cluster GPS coordinates and geospatial covariates, are third-party data publicly available from UNICEF at https://mics.unicef.org, subject to registration and the MICS terms of use. Other researchers can access them in the same way as the authors.District boundaries are openly available from geoBoundaries (https://www.geoboundaries.org).

https://mics.unicef.org/surveys

https://www.geoboundaries.org

## Acknowledgments

The authors thank the Bangladesh Bureau of Statistics and UNICEF Bangladesh for making the Multiple Indicator Cluster Survey microdata publicly available.

## Supporting information

**S1 Appendix. Detailed methods and results.** Full text of the Methods and Results summarized in the main text, including missing-data codes, bootstrap details, fixed-list intervals, optimism and sensitivity analyses, spatial neighbors and environmental data sources.

**S1 Table. District selection analyses.** (A) All rules and sensitivity analyses at 10%, 20% and 30% coverage. (B) Paired differences between rules. (C) Lists held fixed at the full-sample selection. (D) Bootstrap optimism. (E) Stunting hotspot list. (F) District prevalence and ranks, 2025 and 2019. (G) Districts in each list and selection frequencies.

**S2 Table. District hotspot analyses (secondary).** Hotspots, environmental adjustment and prevalence by fieldwork month.

**S3 Table. Individual-level environmental models (secondary).** Survey-weighted logistic models of six cluster-level environmental and access factors, with all coefficients and goodness of fit of the primary model.

**S1 Fig.**
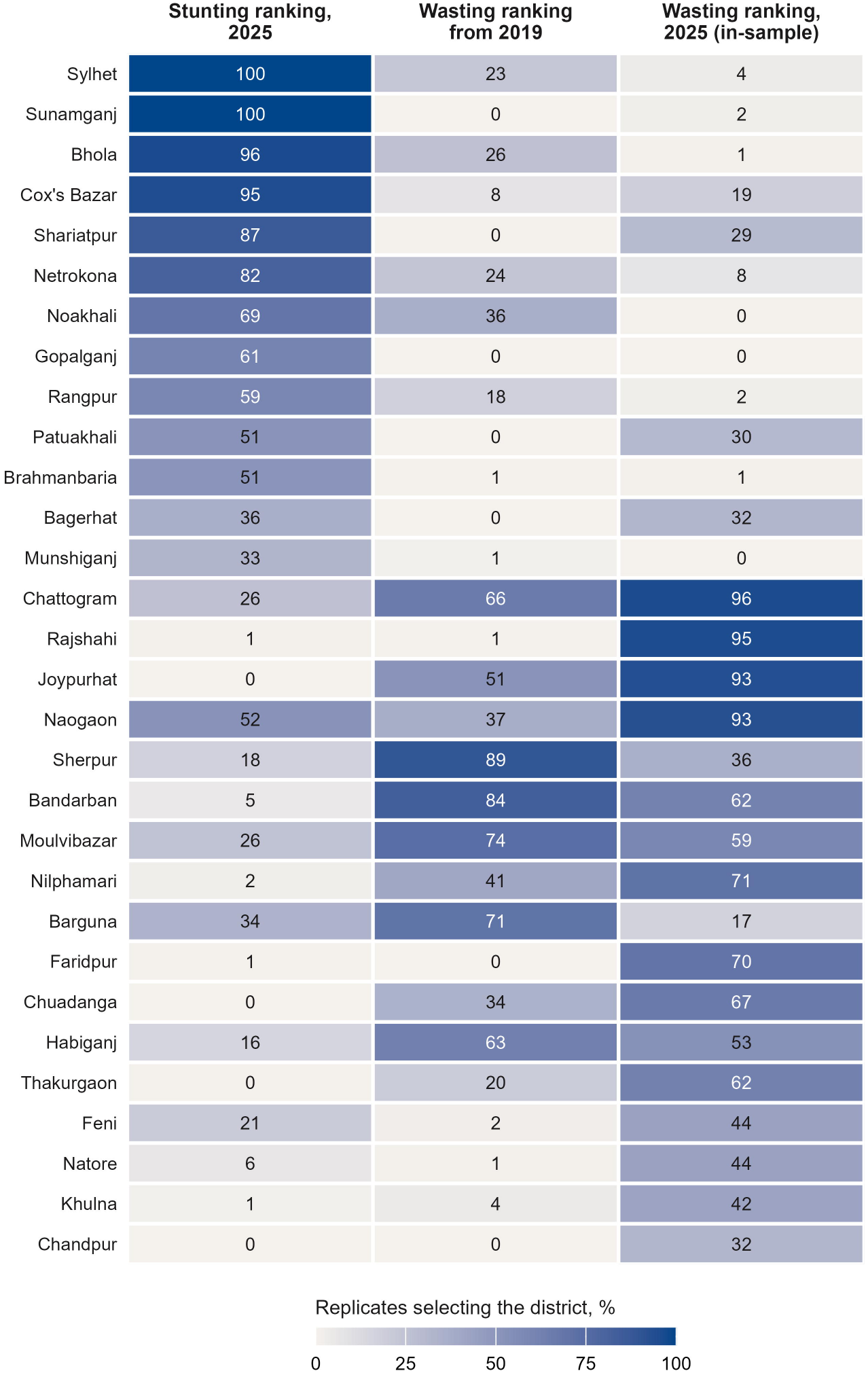
Selection frequency of districts at about 20% coverage. Each cell shows the share of 2,000 bootstrap replicates in which the district was selected by that ranking. The 30 districts shown are those selected most often by the 2025 stunting or wasting ranking. Frequencies describe list stability, not the probability that a district belongs in an ideal list.

**S2 Fig.**
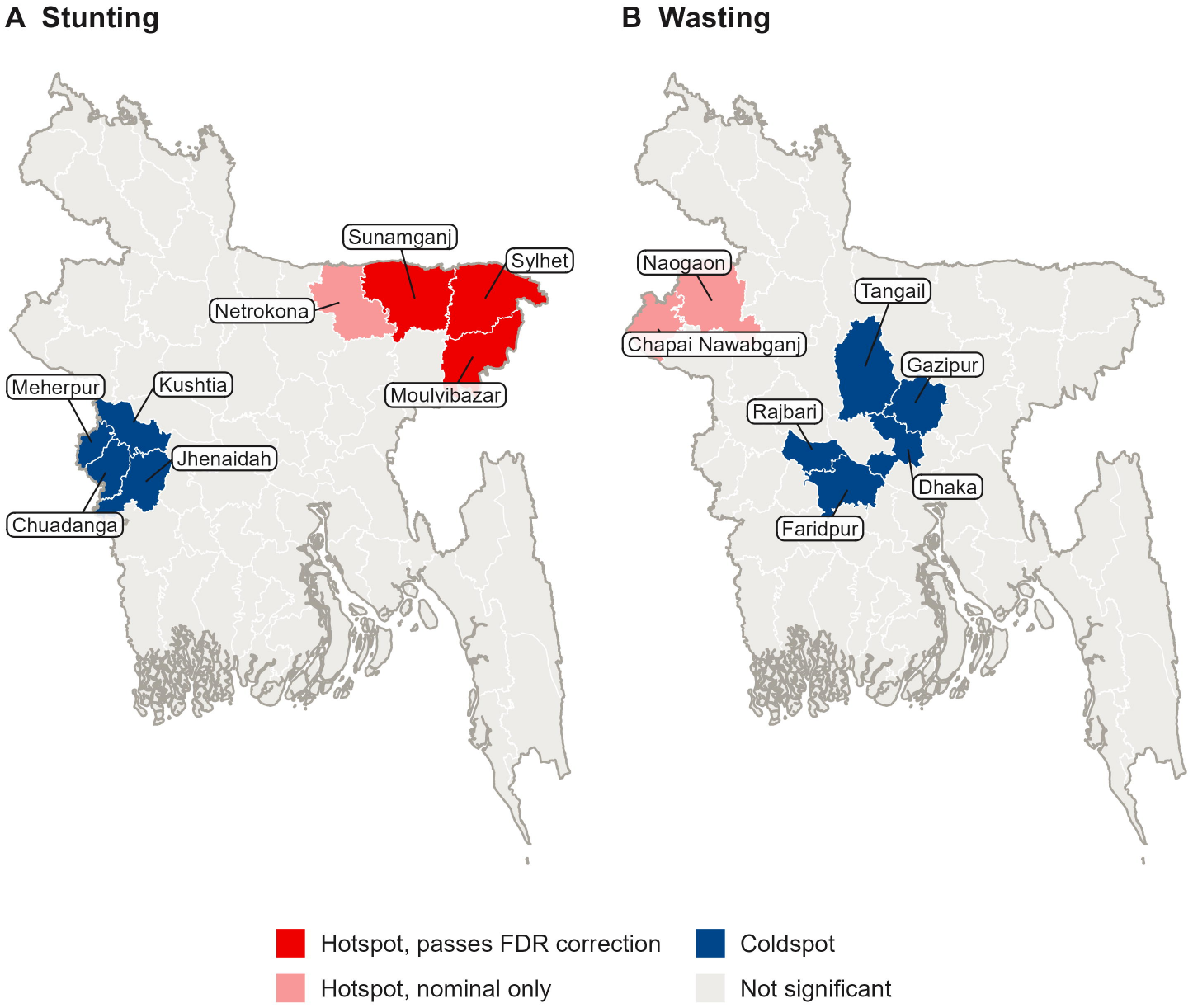
Hotspot and coldspot districts for stunting (A) and wasting (B), Bangladesh MICS 2025 (secondary). Districts are classified by the Getis-Ord Gi* statistic on survey-weighted district prevalence (|z| > 1.96), with five nearest-centroid neighbors. Darker red hotspots also pass Benjamini-Hochberg false discovery rate correction across 64 districts. District boundaries are from geoBoundaries.

**S3 Fig.**
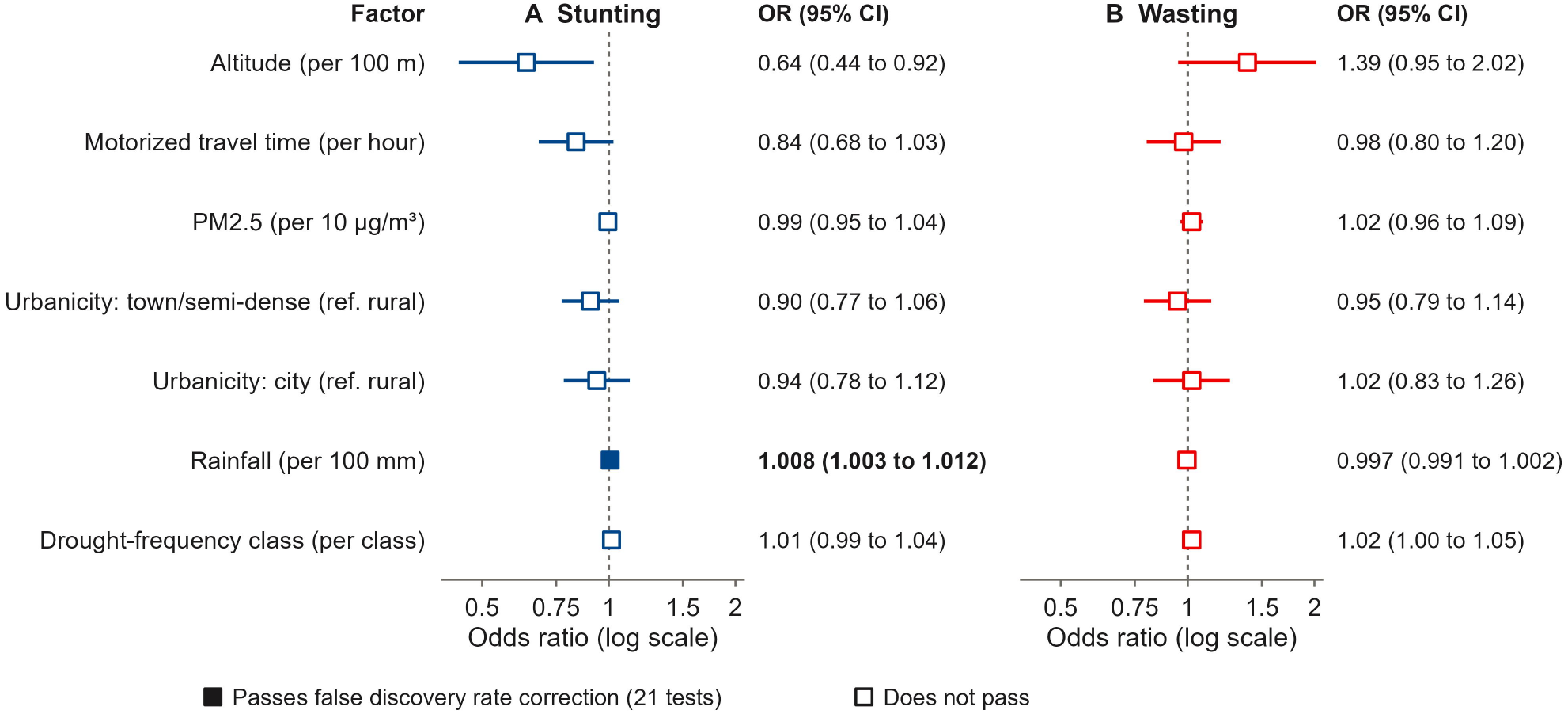
Adjusted associations of cluster-level environmental factors with stunting (A) and wasting (B) (secondary). Odds ratios are from survey-weighted logistic models adjusted for child age group, sex, household wealth quintile, area and division. False discovery rate adjustment covers 21 tests (seven terms, three outcomes); the figure shows two outcomes. Adjusted P values are in S3 Table, panel A.

**S4 Fig.**
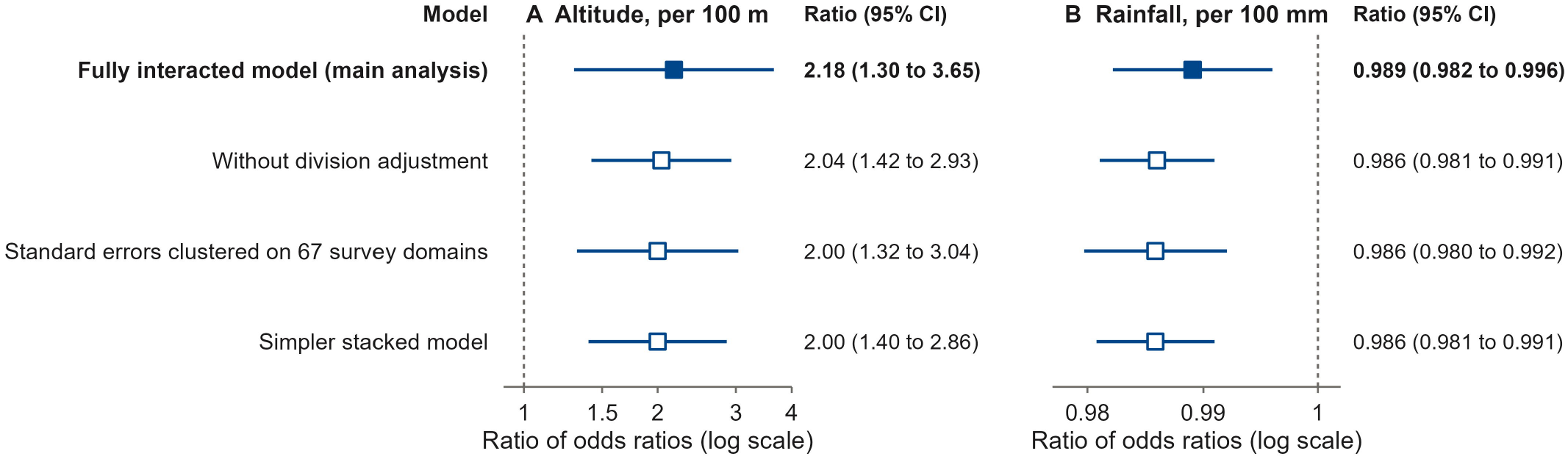
Difference between wasting and stunting in their associations with altitude (A) and rainfall (B) (secondary). Each row gives the ratio of odds ratios, wasting relative to stunting, from one model in S3 Table, panel D. A ratio of 1 means no difference. The altitude association was not linear (S3 Table, panel C).

**S1 Checklist. STROBE checklist for cross-sectional studies.**

