## Supplementary material for "Geographic prioritization of child nutrition services in Bangladesh: comparing stunting and wasting indicators using the 2025 Multiple Indicator Cluster Survey": S1_Appendix.docx

### **S1 Appendix. Detailed methods and results**

This appendix gives the full Methods and Results text that the main text summarizes. Bracketed numbers refer to the main-text reference list, except those marked A, which are listed at the end of this appendix. Tables and figures are those of the main text and supplement.

### **Methods**

#### **Study design and data**

This is a cross-sectional analysis of the Bangladesh MICS 2025, conducted by the Bangladesh Bureau of Statistics with UNICEF Bangladesh [5]. The survey used a stratified two-stage cluster sample of enumeration areas and households. Fieldwork ran from 24 February to 30 June 2025, and the household response rate was 99.6% [5]. For the temporal check, we used the Bangladesh MICS 2019, which had a similar design and the same anthropometric indicators [17]. Both surveys are nationally representative and provide child weights, strata and primary sampling units (PSUs).

#### **Population and outcomes**

The population was all children under five listed in the 2025 survey. We included every eligible child, so no sample size calculation was needed. Wasting was defined as a weight-for-height z-score below −2, and stunting as a height-for-age z-score below −2 (2006 World Health Organization child growth standards). We used the z-scores released by MICS against these standards (variables HAZ2 and WHZ2). This is the survey definition of wasting and does not use mid-upper arm circumference. A z-score was valid if the survey had calculated it and had not assigned an exclusion code. MICS assigns 99.99 to a missing measurement and 99.98 to a value flagged as implausible under WHO criteria. It assigns 99.97 to other values that could not be scored. We confirmed the 99.98 flags by recomputing z-scores with the WHO anthro R package. The common denominator for all coverage measures was children with a valid wasting measurement. Children with a valid stunting measurement provided the stunting prevalence used for ranking. The 2019 data were prepared with the same rules.

#### **Districts**

Bangladesh has 64 districts. MICS 2025 reported 67 survey domains, because the Dhaka North, Dhaka South and Chattogram city corporations were reported separately. We merged each city corporation into its parent district, giving 64 districts in both surveys. District names were matched to geoBoundaries [18], and every child in both surveys was assigned a district.

#### **Selection rules**

We compared four rules for choosing districts. Rules A and B rank districts from the highest to the lowest prevalence and add them in rank order. Rule A ranked districts by stunting prevalence in 2025. This is the strategy of interest. Rule B ranked districts by wasting prevalence in 2025. It uses the same data that are used to judge it, so it is an in-sample benchmark expected to look better than it would in practice. Rule C was population-proportional allocation. It selects the same fraction c of children in every district, so the share of wasted children included is c by construction. It does not rank districts by population size. Rule D was the fixed list of four nominal stunting hotspot districts (Moulvibazar, Netrokona, Sunamganj and Sylhet). Our Getis-Ord analysis of the same 2025 data identified them, and three passed false discovery rate correction (see Secondary spatial and environmental analyses; S2 Table).

As secondary rules, we ranked districts by the number of stunted children and by 2019 stunting prevalence. Prevalence estimates had no exact ties.

#### **Coverage and burden measures**

Coverage is the weighted share of children with a valid wasting measurement who live in the selected districts. Burden capture is the weighted share of wasted children who live there. Enrichment is burden capture divided by coverage. An enrichment of 1 means that the list includes wasted children exactly in proportion to their share of all children, as under rule C. An enrichment of 1.2 means 20% more wasted children than population-proportional allocation at the same coverage. We also report burden capture minus coverage in percentage points. Survey weights are normalized, so we report shares of children and not national counts.

We used analytical coverage scenarios of about 10%, 20% and 30% of children. These are reference points for comparison, not recommended program sizes. Whole districts rarely add up to an exact target. We therefore added districts until coverage first reached the target, and we report the achieved coverage of each list. For comparison at exactly the same coverage, we also interpolated along each ranking. This assumes that only part of the last district is included, with the same wasting prevalence as the whole district. Interpolated results are labeled as coverage-matched and are shown beside the whole-district lists. Coverage curves show burden capture against coverage over all possible list sizes.

#### **Survey uncertainty**

We estimated uncertainty with a stratified rescaling bootstrap for complex surveys [19]. In each replicate, we drew with replacement one fewer primary sampling unit (PSU) than each stratum contained, and rescaled the weights. All 131 strata in 2025 and 128 in 2019 had at least two PSUs. Three 2025 PSUs and four 2019 PSUs had completed household interviews but no listed children. A rerun that included them with zero child totals gave identical point estimates. Interval limits changed by 0.1 percentage points or less for the primary comparison, by at most 0.4 points elsewhere and by at most 0.02 for enrichment. Each of 2,000 replicates repeated the full process: district prevalence, ranking, selection, coverage and burden. All rules used the same replicates, so differences between rules are paired. We report percentile 95% confidence intervals (CIs) and the share of replicates in which each district was selected. Selection frequencies describe list stability under resampling, not the probability that a district belongs in an ideal list. For rules ranked and evaluated on the same outcome in the same survey (rule B), we omit percentile intervals for burden capture and enrichment under repeated selection. Ranking by observed prevalence maximizes the observed share by construction, so these intervals lack nominal coverage. Intervals for lists held fixed are reported separately (S1 Table, panel C). For the stunting ranking, we also built a simultaneous 95% band over coverage from 5% to 95%, in steps of one percentage point. Each replicate curve was standardized by its bootstrap mean and standard deviation. The critical value was the 95th percentile of the largest absolute standardized deviation. No replicate produced an undefined district estimate.

All intervals are 95% confidence intervals. These intervals describe the selection procedure. They include the chance that a different set of districts would have been chosen. We also report intervals for lists held fixed at the full-sample selection, which describe uncertainty in burden for that one list. For rule D, we also estimated a fixed-list interval by Taylor linearization. We made no adjustment for multiple comparisons across rules, coverage levels and survey years. Contrasts other than the primary comparison are exploratory.

#### **Selection optimism**

A rule that ranks districts by 2025 wasting and is judged on 2025 wasting will look better than it would in practice. Part of what it captures is sampling noise. Our main treatment of this optimism was a temporal assessment. We ranked districts by wasting prevalence in MICS 2019 and judged the resulting lists against 2025 wasting. The 2019 ordering was applied to the 2025 child population, so lists stopped at 2025 coverage. In each replicate, the 2019 sample was resampled independently of the 2025 sample. This reflects how a planner would use the most recent survey before new data arrive. The six-year gap, the COVID-19 period and the rise in national wasting may have made the 2019 ranking outdated. This check may therefore understate what a current ranking could achieve. As a supporting check, we estimated bootstrap optimism [20]. Each replicate ranking was also applied to the full-sample district shares. Optimism was the mean amount by which a replicate ranking did better in its own replicate than in the full sample. We subtracted it from the in-sample estimate.

#### **Sensitivity analyses**

Wasting changes with season, and fieldwork reached districts in different months. We therefore standardized for month of measurement. For each district, we calculated the number of wasted children expected from its fieldwork timing. In each month, we multiplied the weighted number of children measured in the district by national wasting prevalence in that month. These products were summed over months. Districts were ranked by the ratio of observed to expected wasted children. Each district’s adjusted share of wasted children was its weighted number of children multiplied by this ratio. These values were then rescaled to sum to one across districts. This method assumed that seasonal patterns were the same across districts, and it does not estimate annual burden. Every district had children measured in at least two of the five fieldwork months.

Missing anthropometry could bias district estimates. MICS child weights already adjust for children without a completed questionnaire. Among interviewed children, we distinguished missing measurements (code 99.99), implausible values (99.98) and values that could not be scored (99.97). We described missingness by division, age group, wealth quintile and area with design-based tests. We then reweighted children with a valid z-score by the inverse of the weighted response rate within cells of district by six age groups. Each outcome used its own valid sample, and coverage used all interviewed children. A cell with no valid measurement took its district’s prevalence. Selection analyses were repeated with these weights. Month standardization and reweighting were recomputed in every bootstrap replicate. We did not impute values.

We also ran a reverse analysis: the share of stunted children included by wasting-based selection. This was secondary, because the policy question concerns wasting.

#### **Secondary spatial and environmental analyses**

Secondary analyses described the spatial pattern of each condition. We tested clustering of district prevalence with Moran’s I [21] and identified hotspots with the Getis-Ord Gi* statistic [22], with false discovery rate correction. A hotspot was a district with Gi* z above 1.96, and correction used the Benjamini-Hochberg method at a false discovery rate of 0.05. The stunting hotspots defined rule D. Survey-weighted logistic models related six cluster-level environmental and access factors to each outcome. These models describe patterns and do not inform district selection. Full results are in S2 and S3 Tables and S2 to S4 Figs.

#### **Spatial neighbors**

Each district’s neighbors were the five districts whose centroids lay closest to its own centroid. Centroids came from the geoBoundaries district polygons [18]. Each neighbor received equal weight, so the weights for each district summed to one (row standardization). For Gi*, each district was also included as its own neighbor. The same neighbors were used for Moran’s I, Gi* and the residual analyses (S2 Table).

Distances between centroids were computed from longitude and latitude without map projection. District centroids lie between about 21.5° and 26.3° north. When we recomputed neighbors with great-circle distances, 58 of 64 districts kept the same five neighbors, and 98% of all neighbor links were unchanged.

#### **Environmental and access factors**

The MICS 2025 geospatial covariate file links each cluster to over 150 environmental and access variables [A1]. MICS calculated each value from gridded data within a 2 km (urban) or 5 km (rural) buffer around the displaced cluster location [A1]. It used the mean or, for categorical layers, the most common class.

We chose six factors that were not strongly correlated with each other. These were altitude, motorized travel time to the nearest health facility, fine particulate matter (PM2.5), urbanicity, annual rainfall and drought frequency. We dropped variables that overlapped with these. Night-time lights and population density were correlated with each other (r=0.80). Walking travel time was correlated with motorized travel time (r=0.75), and mean temperature with rainfall (r=−0.73).

Altitude came from a digital elevation model at about 1 km resolution, distributed through the Oak Ridge National Laboratory (ORNL) Spatial Data Access Tool [A1]. Rainfall was the 2020 annual total from the Climate Hazards Center Infrared Precipitation with Stations (CHIRPS) version 3 product, on a 0.05° grid [A2]. PM2.5 was the 2020 annual mean from the Atmospheric Composition Analysis Group estimates, on a 0.01° grid [A3]. Motorized and walking travel times were 2019 estimates from the Malaria Atlas Project [A4]. Urbanicity was the 2025 three-level Degree of Urbanisation class from the Global Human Settlement Layer [A5]. Drought was a decile class of drought frequency for 1980 to 2000 [A6]. Continuous factors were rescaled: altitude per 100 m, travel time per hour, PM2.5 per 10 µg/m³ and rainfall per 100 mm.

### **Results**

#### **Sample and missing data**

Of 24,680 listed children, 23,357 (94.6%) had a completed questionnaire (Table 1). Among them, 1,037 had no valid height-for-age z-score and 1,046 had no valid weight-for-height z-score. Most were missing a measurement (856 and 866). Others had values flagged as implausible (180 and 125), and the values of 1 and 55 children could not be scored. This left 22,320 children for stunting and 22,311 for wasting, with 22,205 measured for both. In the text, counts are unweighted and percentages are survey-weighted. The analysis covered 64 districts, 131 strata and 3,146 PSUs.

Notes on Table 1. Children without a completed questionnaire have zero weight in the released file because MICS weights adjust for questionnaire nonresponse. Counts are unweighted; percentages are weighted unless stated. When z-scores were recomputed with the WHO anthro R package, every checkable 99.98 value met WHO flag criteria (176 of 180 for height-for-age, 125 of 125 for weight-for-height). Design-based tests of height-for-age missingness: division P<0.001, age group P=0.021, wealth quintile P=0.23, urban or rural P=0.78. Missing codes are as assigned by MICS.

Weighted missingness of the height-for-age z-score was 4.6% overall (1,037 of 23,357 interviewed children). It varied by division (design-based P<0.001), from 3.2% in Rajshahi (72 of 2,333) to 7.9% in Mymensingh (104 of 1,430) and 7.0% in Sylhet (114 of 1,655). It also varied by age (P=0.021) and was highest in infants under 6 months (6.2%; 135 of 2,224). It did not vary by wealth quintile (P=0.23) or by urban or rural residence (P=0.78). Missingness of the weight-for-height z-score followed the same pattern (Table 1).

Weighted prevalence was 24.0% for stunting (5,365 of 22,320; 95% CI 23.3 to 24.7) and 12.9% for wasting (2,905 of 22,311; 95% CI 12.4 to 13.4). Concurrent wasting and stunting affected 4.0% (902 of 22,205; 95% CI 3.7 to 4.3). Among wasted children with both measurements, 31.2% (902 of 2,881; 95% CI 29.2 to 33.2) were also stunted (Table 1).

#### **District distributions**

District stunting prevalence ranged from 17.0% in Kushtia (51 of 306) to 37.3% in Sylhet (145 of 389). District wasting prevalence ranged from 6.3% in Manikganj (18 of 268) to 20.0% in Naogaon (40 of 211) (Fig 1). District stunting and wasting prevalence showed no linear association (Pearson r −0.01 and Spearman 0.06 across 64 districts, given descriptively). The highest stunting was in the northeast (Sylhet, Sunamganj and Netrokona) and in southern districts (Bhola, Shariatpur, Cox’s Bazar and Noakhali). The highest wasting was mostly in the northwest and west (Naogaon, Rajshahi, Joypurhat and Chuadanga), and in Chattogram and Faridpur (Fig 1).

District wasting estimates were imprecise. Districts had a median of 324 children with a valid wasting measurement (range 211 to 832; S1 Table, panel F). The median width of their 95% confidence intervals was 7.5 percentage points (range 4.0 to 12.0). Between 2019 and 2025, district wasting prevalence correlated weakly (r 0.32), and district stunting prevalence correlated moderately (r 0.45; S1 Table, panel F).

#### **Primary comparison**

At the three coverage levels, districts ranked by stunting included wasted children at about their population share (Table 2, Fig 2). The first 7 districts by stunting held 12.3% of children (2,866 of 22,311) and 11.7% of wasted children (359 of 2,905). Their enrichment was 0.95 (95% CI 0.84 to 1.14). The first 13 districts held 20.1% of children (4,726) and 19.6% of wasted children (595), with enrichment 0.98 (0.89 to 1.13). The first 19 districts held 30.4% of children (7,036) and 31.8% of wasted children (921), with enrichment 1.04 (0.93 to 1.11).

Coverage-matched estimates gave the same picture (Table 2). At exactly 10%, 20% and 30% coverage, stunting-ranked districts included 9.7%, 19.5% and 31.3% of wasted children. These differed from population-proportional allocation by −0.3 (95% CI −1.6 to 1.2), −0.5 (−2.2 to 2.6) and 1.3 (−2.3 to 3.3) percentage points. At these three levels, the intervals excluded gains or losses larger than about 3.3 percentage points. Across the whole curve, the simultaneous band was wider: −4.2 to 3.2 points at 20% coverage and −3.2 to 5.8 at 30% (Fig 2).

Districts ranked by 2025 wasting included 14.1%, 26.3% and 37.4% of wasted children at the same three coverage levels (enrichment 1.41, 1.32 and 1.25). The stunting ranking included 4.4, 6.8 and 6.2 percentage points fewer wasted children (S1 Table, panel B). Ranking by observed wasting maximizes this share by construction, so these in-sample figures have no intervals. They overstate what a wasting ranking would achieve, as the next sections show.

The four stunting hotspot districts held 5.9% of children (1,471 of 22,311; 95% CI 5.5 to 6.3). They held 5.9% of wasted children (197 of 2,905; 95% CI 5.0 to 6.8). Their enrichment was 1.00 (0.85 to 1.15) by bootstrap and 1.00 (0.85 to 1.15) by linearization (Table 2; S1 Table, panel E).

#### **Uncertainty and stability of district lists**

District lists changed between replicates. To reach 10% coverage, the stunting ranking needed 4 to 11 districts across replicates, and the wasting ranking 4 to 12 (S1 Table, panel A). Sylhet and Sunamganj entered the 10% stunting list in 99% or more of the 2,000 replicates, Bhola in 78% and Netrokona in 50%. No district entered the 10% wasting list in more than 83% of replicates (Rajshahi 83%, Naogaon 81%, Chattogram and Joypurhat 73%; S1 Table, panel G). S1 Fig shows selection frequencies at about 20% coverage. The stunting and wasting lists shared no district at 10% coverage, one at 20% and eight at 30% (S1 Table, panel G).

Intervals for lists held fixed were narrower than those for the full procedure. For the stunting list at 20% coverage, the fixed-list enrichment was 0.98 (0.90 to 1.05), against 0.98 (0.89 to 1.13) when selection was repeated (S1 Table, panel C). The share of replicates in which stunting-ranked districts included fewer wasted children than population-proportional allocation was 65% at 10%, 55% at 20% and 39% at 30% coverage (S1 Table, panel A).

#### **Selection optimism and the temporal assessment**

At matched coverage, 2019 wasting rankings judged against 2025 data included 10.6% (95% CI 9.0 to 12.7), 22.9% (19.0 to 24.2) and 33.1% (29.2 to 34.7) of wasted children. Their coverage-matched enrichments were 1.06 (0.90 to 1.27), 1.15 (0.95 to 1.21) and 1.10 (0.97 to 1.16). These were lower than the in-sample 2025 values, and every interval included 1. Paired differences (2025 stunting ranking minus 2019 wasting ranking) were −0.8 (−3.3 to 1.3), −3.4 (−5.2 to 1.9) and −1.8 (−5.5 to 2.1) percentage points (S1 Table, panel B).

When both rankings came from 2019, so that both indicators had the same information date, the stunting ranking gave enrichments of 0.91, 0.95 and 0.90 (S1 Table, panel A). Its paired differences from the 2019 wasting ranking were −1.5 (95% CI −3.7 to 1.1), −3.9 (−6.5 to 0.4) and −6.0 (−8.0 to −0.5) percentage points (panel B). This comparison is exploratory.

Bootstrap optimism gave a similar warning. The estimated optimism of the in-sample wasting ranking was 1.4, 2.1 and 2.7 percentage points, giving corrected enrichments of 1.27, 1.21 and 1.16 (S1 Table, panel D). At 20% coverage, the wasting ranking therefore included 26.3% of wasted children in-sample and 24.3% after optimism correction. The 2019 wasting ranking included 22.9%. Estimated optimism for the stunting ranking was 0.15 percentage points or less. For the in-sample wasting ranking at 30% coverage, the bootstrap percentile range (37.5% to 40.6%) lay above the observed share (37.4%).

#### **Sensitivity analyses**

Month standardization changed the results little. Every district had children measured in at least two months, and 40 of 64 districts had children measured in four or five. Coverage-matched enrichment of the stunting ranking was 0.99, 0.98 and 1.04 after standardization. The hotspot list gave 1.01 (0.86 to 1.16). Response reweighting gave 0.98, 0.97 and 1.04 for the stunting ranking and 1.00 (0.86 to 1.16) for the hotspot list (S1 Table, panels A and E).

Ranking by the number of stunted children gave enrichments of 1.07 (0.79 to 1.25), 0.96 (0.87 to 1.06) and 0.95 (0.88 to 1.01). In the reverse analysis, districts ranked by wasting included stunted children in proportion to their population share (enrichment 1.01, 0.99 and 0.99). Full results are in S1 Table, panel A.

#### **Secondary spatial and environmental results**

Stunting clustered across districts (Moran’s I 0.21, P=0.003), with four nominal hotspot districts in the northeast, three of which passed false discovery rate correction (S2 Fig). Wasting showed two nominal hotspot districts, Naogaon and Chapai Nawabganj, and none passed correction. District clustering of wasting was not detected (I 0.05, P=0.17). After adjustment for six environmental and access factors, stunting clustering was no longer detected. The role of these factors could not be separated from region (S2 Table, panels B and C). Full results are in S2 and S3 Tables.
