## Supplementary material for "Geographic prioritization of child nutrition services in Bangladesh: comparing stunting and wasting indicators using the 2025 Multiple Indicator Cluster Survey": S1_Table.docx

### **S1 Table. District selection analyses, Bangladesh MICS 2025**

All intervals are percentile 95% intervals from 2,000 paired stratified bootstrap replicates (in each stratum, one fewer primary sampling unit than the stratum contained was drawn, and weights were rescaled). Each replicate repeated district estimation, ranking and selection. The 2019 survey was resampled independently. Three 2025 PSUs and four 2019 PSUs with completed household interviews but no listed children were not in the resampling frame. A rerun that included them with zero child totals gave identical point estimates, and interval limits changed by at most 0.4 percentage points (0.02 for enrichment). Coverage and burden shares use children with a valid weight-for-height z-score; reverse analyses use children with a valid height-for-age z-score. Enrichment is the share of the outcome divided by the share of children in the selected districts. In-sample rules rank on the same data that judge them, so they maximize the observed share by construction. Their percentile intervals lack nominal coverage, so in-sample rows omit intervals for burden capture and enrichment when selection is repeated. Intervals for achieved child-population coverage are kept, and panel C separately describes uncertainty for lists held fixed. Rankings from the 2019 survey were applied to 2025 child-population shares, so their lists stop at 2025 coverage.

#### **Panel A. All rules and sensitivity analyses**

| Rule | Scenario | Districts, n (range across replicates) | Children in selected districts, % (95% CI) | Outcome in selected districts, % (95% CI) | Enrichment (95% CI) | Coverage-matched outcome share, % (95% CI) | Coverage-matched enrichment (95% CI) | Replicates below proportional, % |
| --- | --- | --- | --- | --- | --- | --- | --- | --- |
| A Stunting prevalence, 2025 | 10% | 7 (4 to 11) | 12.3 (10.0 to 13.8) | 11.7 (8.9 to 14.3) | 0.95 (0.84 to 1.14) | 9.7 (8.4 to 11.2) | 0.97 (0.84 to 1.12) | 65.0 |
| A Stunting prevalence, 2025 | 20% | 13 (8 to 17) | 20.1 (20.0 to 24.7) | 19.6 (18.3 to 26.2) | 0.98 (0.89 to 1.13) | 19.5 (17.8 to 22.6) | 0.98 (0.89 to 1.13) | 55.0 |
| A Stunting prevalence, 2025 | 30% | 19 (12 to 26) | 30.4 (30.1 to 34.7) | 31.8 (28.4 to 36.6) | 1.04 (0.93 to 1.11) | 31.3 (27.7 to 33.3) | 1.04 (0.92 to 1.11) | 39.2 |
| Number of stunted children, 2025 | 10% | 2 (2 to 3) | 12.2 (10.4 to 16.4) | 12.3 (9.5 to 16.2) | 1.01 (0.83 to 1.19) | 10.7 (7.9 to 12.5) | 1.07 (0.79 to 1.25) | 41.4 |
| Number of stunted children, 2025 | 20% | 4 (4 to 5) | 20.5 (20.1 to 23.8) | 19.7 (18.1 to 23.3) | 0.96 (0.87 to 1.05) | 19.3 (17.5 to 21.1) | 0.96 (0.87 to 1.06) | 78.6 |
| Number of stunted children, 2025 | 30% | 8 (7 to 10) | 30.0 (30.1 to 32.9) | 28.6 (27.0 to 32.0) | 0.95 (0.88 to 1.01) | 28.6 (26.3 to 30.4) | 0.95 (0.88 to 1.01) | 93.6 |
| B Wasting prevalence, 2025 (in-sample) | 10% | 6 (4 to 12) | 11.1 (10.0 to 14.9) | 15.6 | 1.40 | 14.1 | 1.41 | Not shown |
| B Wasting prevalence, 2025 (in-sample) | 20% | 14 (10 to 19) | 20.4 (20.0 to 23.5) | 26.8 | 1.31 | 26.3 | 1.32 | Not shown |
| B Wasting prevalence, 2025 (in-sample) | 30% | 23 (16 to 26) | 32.0 (30.0 to 32.6) | 39.6 | 1.24 | 37.4 | 1.25 | Not shown |
| Stunting prevalence, 2019 survey | 10% | 7 (3 to 10) | 12.0 (10.0 to 15.7) | 11.1 (8.7 to 13.6) | 0.92 (0.79 to 1.07) | 9.1 (8.0 to 10.8) | 0.91 (0.80 to 1.08) | 78.8 |
| Stunting prevalence, 2019 survey | 20% | 12 (7 to 15) | 24.4 (20.1 to 26.2) | 22.2 (17.2 to 24.1) | 0.91 (0.82 to 1.03) | 19.1 (16.5 to 20.7) | 0.95 (0.82 to 1.04) | 88.1 |
| Stunting prevalence, 2019 survey | 30% | 14 (12 to 21) | 30.3 (30.0 to 35.6) | 27.4 (26.1 to 33.2) | 0.90 (0.85 to 1.00) | 27.1 (25.5 to 30.2) | 0.90 (0.85 to 1.01) | 96.2 |
| Wasting prevalence, 2019 survey | 10% | 9 (4 to 12) | 10.5 (10.0 to 14.8) | 11.1 (9.4 to 17.4) | 1.06 (0.90 to 1.27) | 10.6 (9.0 to 12.7) | 1.06 (0.90 to 1.27) | 24.6 |
| Wasting prevalence, 2019 survey | 20% | 14 (8 to 19) | 20.2 (20.0 to 24.8) | 23.2 (19.6 to 27.5) | 1.15 (0.95 to 1.21) | 22.9 (19.0 to 24.2) | 1.15 (0.95 to 1.21) | 12.0 |
| Wasting prevalence, 2019 survey | 30% | 21 (14 to 26) | 31.5 (30.0 to 34.9) | 34.6 (29.9 to 37.2) | 1.10 (0.97 to 1.15) | 33.1 (29.2 to 34.7) | 1.10 (0.97 to 1.16) | 7.6 |
| A, month-standardized | 10% | 7 (4 to 11) | 12.3 (10.0 to 13.8) | 11.8 (8.9 to 14.2) | 0.96 (0.84 to 1.15) | 9.9 (8.4 to 11.3) | 0.99 (0.84 to 1.13) | 60.3 |
| A, month-standardized | 20% | 13 (8 to 17) | 20.1 (20.0 to 24.7) | 19.6 (18.4 to 26.2) | 0.98 (0.90 to 1.14) | 19.6 (17.8 to 22.6) | 0.98 (0.89 to 1.13) | 51.8 |
| A, month-standardized | 30% | 19 (12 to 26) | 30.4 (30.1 to 34.7) | 31.8 (28.5 to 36.5) | 1.04 (0.93 to 1.11) | 31.3 (27.8 to 33.3) | 1.04 (0.93 to 1.11) | 37.0 |
| B, month-standardized (in-sample) | 10% | 5 (4 to 12) | 10.3 (10.0 to 14.9) | 14.3 | 1.39 | 13.9 | 1.39 | Not shown |
| B, month-standardized (in-sample) | 20% | 14 (10 to 19) | 20.1 (20.0 to 23.4) | 26.0 | 1.29 | 25.9 | 1.30 | Not shown |
| B, month-standardized (in-sample) | 30% | 22 (16 to 26) | 30.5 (30.0 to 33.1) | 37.5 | 1.23 | 37.0 | 1.23 | Not shown |
| A, response-weighted | 10% | 7 (4 to 10) | 12.5 (10.0 to 13.6) | 11.9 (8.8 to 14.1) | 0.95 (0.84 to 1.13) | 9.8 (8.4 to 11.2) | 0.98 (0.84 to 1.12) | 66.0 |
| A, response-weighted | 20% | 13 (8 to 18) | 20.3 (20.0 to 24.8) | 19.8 (18.4 to 26.5) | 0.97 (0.89 to 1.13) | 19.5 (17.7 to 22.5) | 0.97 (0.89 to 1.12) | 57.3 |
| A, response-weighted | 30% | 19 (12 to 25) | 30.8 (30.0 to 34.5) | 32.1 (28.3 to 36.3) | 1.04 (0.92 to 1.11) | 31.2 (27.6 to 33.2) | 1.04 (0.92 to 1.11) | 42.8 |
| B, response-weighted (in-sample) | 10% | 6 (4 to 12) | 10.9 (10.0 to 15.0) | 15.2 | 1.39 | 14.0 | 1.40 | Not shown |
| B, response-weighted (in-sample) | 20% | 14 (10 to 19) | 20.2 (20.0 to 23.5) | 26.5 | 1.31 | 26.3 | 1.31 | Not shown |
| B, response-weighted (in-sample) | 30% | 21 (16 to 26) | 30.1 (30.0 to 32.9) | 37.5 | 1.25 | 37.4 | 1.25 | Not shown |
| Reverse: B ranking, share of stunted children | 10% | 6 (4 to 12) | 11.1 (10.0 to 14.9) | 11.1 (9.4 to 15.1) | 1.00 (0.90 to 1.13) | 10.1 (8.9 to 11.3) | 1.01 (0.89 to 1.13) | 50.0 |
| Reverse: B ranking, share of stunted children | 20% | 14 (10 to 19) | 20.4 (20.0 to 23.5) | 20.2 (19.1 to 23.8) | 0.99 (0.93 to 1.09) | 19.9 (18.6 to 21.7) | 0.99 (0.93 to 1.09) | 45.6 |
| Reverse: B ranking, share of stunted children | 30% | 23 (17 to 26) | 32.0 (30.0 to 32.7) | 32.3 (29.1 to 33.8) | 1.01 (0.95 to 1.07) | 29.7 (28.6 to 32.1) | 0.99 (0.95 to 1.07) | 40.5 |
| Reverse comparator: A ranking, share of stunted children (in-sample) | 10% | 7 (4 to 11) | 12.3 (10.0 to 13.7) | 16.4 | 1.33 | 13.7 | 1.37 | Not shown |
| Reverse comparator: A ranking, share of stunted children (in-sample) | 20% | 13 (8 to 17) | 20.1 (20.0 to 24.7) | 25.4 | 1.26 | 25.3 | 1.26 | Not shown |
| Reverse comparator: A ranking, share of stunted children (in-sample) | 30% | 19 (12 to 26) | 30.4 (30.1 to 34.6) | 36.7 | 1.20 | 36.2 | 1.21 | Not shown |

Month-standardized rules rank districts by the ratio of observed to expected wasted children. Expected wasted children were calculated month by month, as the weighted number of children measured in the district times national wasting prevalence in that month, and summed. Each district’s adjusted share of wasted children is its weighted number of children multiplied by this ratio, rescaled to sum to one across districts. Response-weighted rules reweight children with a valid z-score by the inverse weighted response rate in cells of district by six age groups. Each outcome uses its own valid sample, coverage uses all interviewed children, and a cell with no valid measurement takes its district’s prevalence. Both adjustments are recomputed in every replicate. Replicates below proportional is the share of replicates in which the rule included fewer wasted children than population-proportional allocation.

#### **Panel B. Paired differences between rules at matched coverage**

| Scenario | Contrast | Difference, percentage points (95% CI) |
| --- | --- | --- |
| 10% | A Stunting prevalence, 2025 minus B Wasting prevalence, 2025 (in-sample) | −4.4 |
| 10% | A Stunting prevalence, 2025 minus Wasting prevalence, 2019 survey | −0.8 (−3.3 to 1.3) |
| 10% | A Stunting prevalence, 2025 minus Stunting prevalence, 2019 survey | 0.6 (−1.3 to 2.2) |
| 10% | Number of stunted children, 2025 minus A Stunting prevalence, 2025 | 1.0 (−2.2 to 3.2) |
| 10% | Stunting prevalence, 2019 survey minus Wasting prevalence, 2019 survey | −1.5 (−3.7 to 1.1) |
| 10% | A, month-standardized minus B, month-standardized (in-sample) | −4.0 |
| 10% | A, response-weighted minus B, response-weighted (in-sample) | −4.2 |
| 20% | A Stunting prevalence, 2025 minus B Wasting prevalence, 2025 (in-sample) | −6.8 |
| 20% | A Stunting prevalence, 2025 minus Wasting prevalence, 2019 survey | −3.4 (−5.2 to 1.9) |
| 20% | A Stunting prevalence, 2025 minus Stunting prevalence, 2019 survey | 0.5 (−1.4 to 4.5) |
| 20% | Number of stunted children, 2025 minus A Stunting prevalence, 2025 | −0.2 (−3.8 to 2.3) |
| 20% | Stunting prevalence, 2019 survey minus Wasting prevalence, 2019 survey | −3.9 (−6.5 to 0.4) |
| 20% | A, month-standardized minus B, month-standardized (in-sample) | −6.3 |
| 20% | A, response-weighted minus B, response-weighted (in-sample) | −6.8 |
| 30% | A Stunting prevalence, 2025 minus B Wasting prevalence, 2025 (in-sample) | −6.2 |
| 30% | A Stunting prevalence, 2025 minus Wasting prevalence, 2019 survey | −1.8 (−5.5 to 2.1) |
| 30% | A Stunting prevalence, 2025 minus Stunting prevalence, 2019 survey | 4.1 (−0.8 to 6.4) |
| 30% | Number of stunted children, 2025 minus A Stunting prevalence, 2025 | −2.7 (−5.3 to 1.3) |
| 30% | Stunting prevalence, 2019 survey minus Wasting prevalence, 2019 survey | −6.0 (−8.0 to −0.5) |
| 30% | A, month-standardized minus B, month-standardized (in-sample) | −5.7 |
| 30% | A, response-weighted minus B, response-weighted (in-sample) | −6.2 |

All contrasts are exploratory and were not prespecified. The contrast of the two 2019 rankings gives both indicators the same information date.

#### **Panel C. Lists held fixed at the full-sample selection**

These intervals describe uncertainty in burden for one fixed list. They ignore uncertainty in which districts are chosen, including 2019 sampling uncertainty for 2019 rankings. A list chosen and judged on the same data is not independently validated by holding it fixed.

| Rule | Scenario | Wasted children in list, % (95% CI) | Enrichment (95% CI) |
| --- | --- | --- | --- |
| A Stunting prevalence, 2025 | 10% | 11.7 (10.3 to 13.0) | 0.95 (0.84 to 1.05) |
| A Stunting prevalence, 2025 | 20% | 19.6 (17.8 to 21.3) | 0.98 (0.90 to 1.05) |
| A Stunting prevalence, 2025 | 30% | 31.8 (29.7 to 33.8) | 1.04 (0.98 to 1.10) |
| Number of stunted children, 2025 | 10% | 12.3 (10.5 to 14.0) | 1.01 (0.88 to 1.13) |
| Number of stunted children, 2025 | 20% | 19.7 (17.4 to 21.9) | 0.96 (0.87 to 1.05) |
| Number of stunted children, 2025 | 30% | 28.6 (26.1 to 30.8) | 0.95 (0.88 to 1.02) |
| B Wasting prevalence, 2025 (in-sample) | 10% | 15.6 (13.9 to 17.3) | 1.40 (1.27 to 1.53) |
| B Wasting prevalence, 2025 (in-sample) | 20% | 26.8 (24.9 to 28.8) | 1.31 (1.23 to 1.40) |
| B Wasting prevalence, 2025 (in-sample) | 30% | 39.6 (37.4 to 41.7) | 1.24 (1.18 to 1.30) |
| Stunting prevalence, 2019 survey | 10% | 11.1 (9.8 to 12.4) | 0.92 (0.82 to 1.02) |
| Stunting prevalence, 2019 survey | 20% | 22.2 (20.3 to 24.2) | 0.91 (0.84 to 0.98) |
| Stunting prevalence, 2019 survey | 30% | 27.4 (25.3 to 29.4) | 0.90 (0.84 to 0.97) |
| Wasting prevalence, 2019 survey | 10% | 11.1 (10.0 to 12.3) | 1.06 (0.96 to 1.17) |
| Wasting prevalence, 2019 survey | 20% | 23.2 (21.4 to 25.0) | 1.15 (1.07 to 1.23) |
| Wasting prevalence, 2019 survey | 30% | 34.6 (32.6 to 36.6) | 1.10 (1.04 to 1.16) |

#### **Panel D. Bootstrap optimism at matched coverage**

Optimism is the mean, over replicates, of the replicate ranking’s share of wasted children in its own replicate minus its share when applied to the full-sample district shares. The corrected share is the apparent share minus optimism.

| Rule | Scenario | Apparent share, % | Optimism, percentage points | Corrected share, % | Corrected enrichment |
| --- | --- | --- | --- | --- | --- |
| A Stunting prevalence, 2025 | 10% | 9.7 | −0.01 | 9.8 | 0.98 |
| A Stunting prevalence, 2025 | 20% | 19.5 | 0.07 | 19.4 | 0.97 |
| A Stunting prevalence, 2025 | 30% | 31.3 | 0.15 | 31.1 | 1.04 |
| B Wasting prevalence, 2025 (in-sample) | 10% | 14.1 | 1.38 | 12.7 | 1.27 |
| B Wasting prevalence, 2025 (in-sample) | 20% | 26.3 | 2.09 | 24.3 | 1.21 |
| B Wasting prevalence, 2025 (in-sample) | 30% | 37.4 | 2.73 | 34.7 | 1.16 |
| Wasting prevalence, 2019 survey | 10% | 10.6 | −0.01 | 10.6 | 1.06 |
| Wasting prevalence, 2019 survey | 20% | 22.9 | −0.02 | 23.0 | 1.15 |
| Wasting prevalence, 2019 survey | 30% | 33.1 | −0.04 | 33.1 | 1.10 |

#### **Panel E. Four-district stunting hotspot list (Moulvibazar, Netrokona, Sunamganj, Sylhet)**

The list comprises the four nominal Getis-Ord Gi* stunting hotspots from our analysis of the same 2025 data (S2 Table, panel A); three passed false discovery rate correction.

| Measure | Estimate | 95% CI |
| --- | --- | --- |
| Children in list, share | 0.059 | 0.055 to 0.063 |
| Wasted children in list, share | 0.059 | 0.050 to 0.068 |
| Enrichment | 0.997 | 0.851 to 1.146 |
| Enrichment, month-standardized | 1.007 | 0.859 to 1.159 |
| Enrichment, response-weighted | 1.004 | 0.858 to 1.155 |

Taylor linearization of the enrichment for the fixed list: 0.997 (0.849 to 1.146).

#### **Panel F. District prevalence, child-population shares and ranks**

| District | Children, stunting sample | Children, wasting sample | Share of children, % | Stunting, % (95% CI) | Wasting, % (95% CI) | Stunting 2019, % | Wasting 2019, % | Month observed to expected wasting ratio | Rank, stunting 2025 | Rank, wasting 2025 | Rank, wasting 2019 |
| --- | --- | --- | --- | --- | --- | --- | --- | --- | --- | --- | --- |
| Bagerhat | 275 | 275 | 0.84 | 26.4 (20.2 to 32.7) | 14.4 (9.9 to 18.8) | 26.8 | 7.4 | 1.06 | 14 | 17 | 54 |
| Bandarban | 408 | 407 | 0.29 | 22.8 (17.1 to 28.5) | 16.3 (11.7 to 21.0) | 29.5 | 14.4 | 1.21 | 36 | 8 | 4 |
| Barguna | 291 | 290 | 0.57 | 26.6 (21.3 to 31.8) | 13.6 (9.6 to 17.6) | 25.7 | 13.1 | 1.10 | 13 | 27 | 7 |
| Barishal | 313 | 312 | 1.43 | 20.6 (16.4 to 24.9) | 14.0 (9.5 to 18.4) | 35.8 | 11.0 | 1.07 | 49 | 22 | 25 |
| Bhola | 401 | 398 | 1.45 | 32.0 (27.1 to 36.9) | 11.6 (8.5 to 14.8) | 38.0 | 11.3 | 0.93 | 3 | 45 | 20 |
| Bogura | 294 | 294 | 2.26 | 23.3 (18.1 to 28.4) | 11.7 (8.2 to 15.3) | 31.5 | 11.4 | 0.94 | 30 | 44 | 19 |
| Brahmanbaria | 405 | 405 | 2.41 | 27.3 (21.5 to 33.0) | 11.4 (8.2 to 14.7) | 26.9 | 7.9 | 0.87 | 12 | 47 | 47 |
| Chandpur | 403 | 401 | 2.09 | 20.4 (16.3 to 24.6) | 14.5 (10.8 to 18.2) | 23.1 | 7.4 | 1.08 | 50 | 16 | 53 |
| Chapai Nawabganj | 317 | 317 | 1.24 | 23.1 (18.9 to 27.3) | 13.1 (10.0 to 16.1) | 25.2 | 4.8 | 1.06 | 31 | 29 | 62 |
| Chattogram | 742 | 739 | 5.45 | 26.1 (22.0 to 30.2) | 17.6 (14.7 to 20.4) | 23.2 | 12.4 | 1.34 | 17 | 4 | 10 |
| Chuadanga | 316 | 316 | 0.84 | 19.3 (14.7 to 23.9) | 16.6 (11.8 to 21.4) | 21.3 | 11.5 | 1.22 | 57 | 5 | 17 |
| Cox’s Bazar | 508 | 510 | 2.32 | 30.6 (26.9 to 34.4) | 13.9 (10.7 to 17.2) | 34.6 | 9.7 | 1.12 | 4 | 23 | 31 |
| Cumilla | 410 | 409 | 4.23 | 25.8 (21.6 to 30.0) | 12.0 (8.9 to 15.2) | 24.4 | 9.3 | 0.93 | 20 | 39 | 33 |
| Dhaka | 835 | 832 | 6.78 | 19.9 (16.5 to 23.4) | 9.3 (7.3 to 11.2) | 33.4 | 9.1 | 0.70 | 54 | 61 | 36 |
| Dinajpur | 320 | 318 | 1.90 | 22.6 (17.3 to 27.8) | 12.9 (8.6 to 17.3) | 24.7 | 11.2 | 1.02 | 38 | 30 | 21 |
| Faridpur | 328 | 329 | 1.36 | 22.0 (17.9 to 26.0) | 16.5 (12.3 to 20.7) | 24.6 | 7.5 | 1.34 | 42 | 6 | 52 |
| Feni | 428 | 426 | 1.26 | 25.8 (21.7 to 30.0) | 15.2 (12.1 to 18.3) | 23.7 | 9.0 | 1.21 | 19 | 13 | 37 |
| Gaibandha | 299 | 300 | 1.49 | 21.7 (16.1 to 27.3) | 13.8 (8.6 to 18.9) | 30.3 | 7.8 | 1.11 | 45 | 25 | 48 |
| Gazipur | 299 | 296 | 3.20 | 18.8 (13.2 to 24.4) | 10.8 (7.1 to 14.4) | 26.1 | 8.9 | 0.87 | 60 | 55 | 38 |
| Gopalganj | 369 | 370 | 0.83 | 28.2 (24.4 to 32.0) | 9.7 (6.4 to 12.9) | 24.8 | 7.2 | 0.77 | 9 | 59 | 56 |
| Habiganj | 401 | 400 | 1.59 | 25.1 (20.6 to 29.6) | 15.6 (11.9 to 19.2) | 34.9 | 12.7 | 1.16 | 23 | 11 | 9 |
| Jamalpur | 342 | 345 | 1.84 | 25.6 (21.0 to 30.2) | 11.9 (8.3 to 15.5) | 33.2 | 11.0 | 0.96 | 21 | 42 | 23 |
| Jashore | 268 | 266 | 1.66 | 23.5 (18.8 to 28.2) | 13.9 (10.1 to 17.7) | 21.1 | 9.3 | 1.12 | 29 | 24 | 34 |
| Jhalokati | 306 | 306 | 0.36 | 22.9 (16.8 to 28.9) | 12.7 (8.5 to 16.9) | 19.9 | 6.5 | 0.94 | 35 | 31 | 61 |
| Jhenaidah | 307 | 311 | 1.26 | 17.2 (12.3 to 22.0) | 11.2 (7.9 to 14.5) | 22.0 | 8.4 | 0.90 | 63 | 49 | 43 |
| Joypurhat | 265 | 265 | 0.55 | 19.3 (14.5 to 24.0) | 18.5 (14.8 to 22.3) | 29.4 | 12.3 | 1.37 | 58 | 3 | 11 |
| Khagrachhari | 361 | 362 | 0.39 | 20.4 (16.3 to 24.5) | 10.3 (7.3 to 13.3) | 27.1 | 4.0 | 0.82 | 51 | 58 | 64 |
| Khulna | 283 | 283 | 1.54 | 21.9 (17.2 to 26.7) | 15.0 (11.2 to 18.8) | 17.2 | 8.7 | 1.14 | 43 | 14 | 39 |
| Kishoreganj | 381 | 381 | 2.16 | 24.4 (19.4 to 29.3) | 12.7 (8.8 to 16.5) | 36.5 | 8.6 | 1.03 | 25 | 32 | 40 |
| Kurigram | 329 | 327 | 1.49 | 24.0 (19.3 to 28.8) | 13.1 (9.6 to 16.7) | 29.3 | 12.7 | 1.06 | 27 | 28 | 8 |
| Kushtia | 306 | 305 | 1.28 | 17.0 (12.6 to 21.3) | 10.9 (7.2 to 14.6) | 15.3 | 10.6 | 0.80 | 64 | 53 | 30 |
| Lakshmipur | 448 | 449 | 1.73 | 18.5 (15.2 to 21.8) | 10.5 (7.1 to 13.8) | 26.2 | 15.5 | 0.84 | 61 | 56 | 1 |
| Lalmonirhat | 378 | 379 | 0.96 | 21.6 (18.1 to 25.2) | 12.3 (8.1 to 16.5) | 30.3 | 12.1 | 0.99 | 46 | 38 | 12 |
| Madaripur | 356 | 355 | 0.84 | 22.3 (16.6 to 27.9) | 12.3 (8.7 to 16.0) | 27.4 | 10.7 | 0.90 | 39 | 37 | 29 |
| Magura | 329 | 330 | 0.66 | 24.6 (19.2 to 30.0) | 11.1 (7.7 to 14.5) | 24.3 | 6.7 | 0.89 | 24 | 50 | 58 |
| Manikganj | 266 | 268 | 0.78 | 26.3 (20.8 to 31.7) | 6.3 (3.3 to 9.4) | 19.5 | 4.8 | 0.51 | 16 | 64 | 63 |
| Meherpur | 249 | 248 | 0.36 | 19.1 (14.3 to 24.0) | 9.2 (5.8 to 12.5) | 15.6 | 7.7 | 0.75 | 59 | 62 | 50 |
| Moulvibazar | 316 | 315 | 1.11 | 26.0 (20.9 to 31.0) | 16.1 (11.3 to 21.0) | 30.3 | 15.0 | 1.29 | 18 | 9 | 3 |
| Munshiganj | 286 | 287 | 0.89 | 26.4 (21.4 to 31.3) | 8.2 (5.5 to 10.8) | 25.7 | 8.1 | 0.65 | 15 | 63 | 45 |
| Mymensingh | 335 | 336 | 3.99 | 23.0 (18.4 to 27.5) | 11.1 (7.9 to 14.2) | 33.2 | 6.7 | 0.90 | 34 | 51 | 59 |
| Naogaon | 208 | 211 | 1.24 | 27.7 (19.9 to 35.4) | 20.0 (14.0 to 26.0) | 25.1 | 11.5 | 1.61 | 11 | 1 | 16 |
| Narail | 368 | 374 | 0.55 | 19.9 (15.8 to 24.0) | 12.4 (8.6 to 16.2) | 27.4 | 8.6 | 0.93 | 55 | 36 | 41 |
| Narayanganj | 304 | 304 | 2.23 | 18.1 (14.0 to 22.1) | 11.9 (8.7 to 15.0) | 24.3 | 6.6 | 0.95 | 62 | 43 | 60 |
| Narsingdi | 367 | 367 | 1.63 | 23.1 (18.9 to 27.2) | 12.5 (9.4 to 15.5) | 28.6 | 6.8 | 0.92 | 32 | 34 | 57 |
| Natore | 275 | 273 | 1.09 | 22.6 (16.5 to 28.7) | 15.2 (11.0 to 19.5) | 18.2 | 8.5 | 1.12 | 37 | 12 | 42 |
| Netrokona | 334 | 330 | 1.22 | 30.1 (25.0 to 35.2) | 12.4 (8.4 to 16.4) | 36.7 | 11.1 | 0.99 | 6 | 35 | 22 |
| Nilphamari | 376 | 376 | 1.41 | 22.1 (16.9 to 27.4) | 16.5 (12.5 to 20.4) | 29.0 | 11.8 | 1.21 | 41 | 7 | 14 |
| Noakhali | 472 | 468 | 2.93 | 28.5 (24.0 to 33.0) | 10.9 (7.7 to 14.0) | 36.6 | 11.7 | 0.81 | 7 | 54 | 15 |
| Pabna | 298 | 296 | 1.72 | 24.0 (17.7 to 30.4) | 11.2 (7.3 to 15.1) | 26.0 | 9.1 | 0.90 | 26 | 48 | 35 |
| Panchagarh | 351 | 350 | 0.70 | 23.0 (18.5 to 27.4) | 12.5 (8.9 to 16.1) | 40.2 | 11.4 | 1.01 | 33 | 33 | 18 |
| Patuakhali | 283 | 282 | 0.87 | 27.8 (21.5 to 34.2) | 14.1 (9.1 to 19.1) | 27.9 | 7.8 | 1.08 | 10 | 20 | 49 |
| Pirojpur | 306 | 306 | 0.68 | 19.5 (15.2 to 23.8) | 11.0 (7.0 to 15.0) | 21.1 | 13.3 | 0.89 | 56 | 52 | 6 |
| Rajbari | 301 | 301 | 0.72 | 21.7 (16.4 to 27.1) | 9.4 (6.2 to 12.6) | 21.8 | 8.0 | 0.77 | 44 | 60 | 46 |
| Rajshahi | 285 | 285 | 1.69 | 20.3 (14.8 to 25.8) | 19.6 (14.6 to 24.6) | 24.4 | 7.3 | 1.44 | 52 | 2 | 55 |
| Rangamati | 326 | 328 | 0.26 | 21.5 (16.1 to 26.8) | 14.0 (9.2 to 18.8) | 36.0 | 9.4 | 1.09 | 47 | 21 | 32 |
| Rangpur | 303 | 302 | 1.87 | 28.3 (21.9 to 34.8) | 11.5 (7.8 to 15.2) | 15.9 | 10.8 | 0.84 | 8 | 46 | 28 |
| Satkhira | 279 | 279 | 1.32 | 20.2 (14.8 to 25.7) | 10.4 (6.6 to 14.2) | 19.9 | 12.0 | 0.82 | 53 | 57 | 13 |
| Shariatpur | 333 | 334 | 0.78 | 30.6 (25.6 to 35.6) | 14.2 (9.6 to 18.9) | 24.4 | 8.3 | 1.17 | 5 | 18 | 44 |
| Sherpur | 315 | 320 | 0.91 | 25.4 (20.9 to 29.8) | 14.8 (10.9 to 18.7) | 27.5 | 15.2 | 1.09 | 22 | 15 | 2 |
| Sirajganj | 319 | 319 | 1.81 | 23.7 (19.0 to 28.3) | 13.6 (9.4 to 17.8) | 27.2 | 10.8 | 1.05 | 28 | 26 | 27 |
| Sunamganj | 435 | 433 | 1.69 | 36.3 (31.0 to 41.6) | 12.0 (8.6 to 15.4) | 44.2 | 7.6 | 0.89 | 2 | 40 | 51 |
| Sylhet | 389 | 393 | 1.89 | 37.3 (32.1 to 42.6) | 12.0 (8.2 to 15.7) | 37.3 | 11.0 | 0.96 | 1 | 41 | 24 |
| Tangail | 264 | 265 | 2.13 | 22.2 (15.8 to 28.7) | 14.2 (10.5 to 17.9) | 19.3 | 14.1 | 1.09 | 40 | 19 | 5 |
| Thakurgaon | 356 | 353 | 0.98 | 21.0 (16.4 to 25.7) | 16.1 (11.7 to 20.4) | 25.9 | 11.0 | 1.17 | 48 | 10 | 26 |

City corporations are merged into their parent districts. Ranks run from 1 (highest prevalence). Share of children is the weighted share of children with a valid weight-for-height z-score; lists add districts in rank order until these shares first reach the target. Coverage-matched estimates include the needed fraction of the last district, at that district’s wasting prevalence.

#### **Panel G. Selection frequency, % of replicates (bold with asterisk = in the full-sample list)**

Frequencies describe how stable each list is under resampling. They are not the probability that a district belongs in an ideal list.

| District | A 10% | A 20% | A 30% | B 10% | B 20% | B 30% | 2019 wasting 10% | 2019 wasting 20% | 2019 wasting 30% |
| --- | --- | --- | --- | --- | --- | --- | --- | --- | --- |
| Bagerhat | 12 | 36 | **55*** | 12 | 32 | **52*** | 0 | 0 | 0 |
| Bandarban | 1 | 5 | 14 | 35 | **62*** | **80*** | **64*** | **84*** | **92*** |
| Barguna | 10 | **34*** | **55*** | 4 | 17 | 37 | **42*** | **71*** | **87*** |
| Barishal | 0 | 0 | 1 | 8 | 23 | **42*** | 9 | 24 | 44 |
| Bhola | **78*** | **96*** | **99*** | 0 | 1 | 5 | 8 | 26 | **50*** |
| Bogura | 1 | 4 | 13 | 0 | 2 | 8 | 17 | 35 | **52*** |
| Brahmanbaria | 20 | **51*** | **70*** | 0 | 1 | 5 | 0 | 1 | 3 |
| Chandpur | 0 | 0 | 1 | 9 | 32 | **57*** | 0 | 0 | 0 |
| Chapai Nawabganj | 0 | 2 | 10 | 1 | 8 | 25 | 0 | 0 | 0 |
| Chattogram | 4 | 26 | **54*** | **73*** | **96*** | **99*** | 27 | **66*** | **88*** |
| Chuadanga | 0 | 0 | 0 | **37*** | **67*** | **82*** | 15 | 34 | **55*** |
| Cox’s Bazar | **67*** | **95*** | **99*** | 4 | 19 | **44*** | 2 | 8 | 18 |
| Cumilla | 4 | 24 | 52 | 0 | 2 | 10 | 0 | 1 | 6 |
| Dhaka | 0 | 0 | 0 | 0 | 0 | 0 | 0 | 2 | 7 |
| Dinajpur | 0 | 4 | 11 | 4 | 12 | 27 | 15 | 29 | **46*** |
| Faridpur | 0 | 1 | 3 | **37*** | **70*** | **86*** | 0 | 0 | 1 |
| Feni | 3 | 21 | **46*** | 12 | **44*** | **73*** | 0 | 2 | 7 |
| Gaibandha | 0 | 4 | 8 | 12 | 28 | 45 | 0 | 0 | 1 |
| Gazipur | 0 | 0 | 0 | 0 | 1 | 4 | 6 | 13 | 20 |
| Gopalganj | 17 | **61*** | **84*** | 0 | 0 | 0 | 0 | 0 | 0 |
| Habiganj | 3 | 16 | 35 | 20 | **53*** | **78*** | **32*** | **63*** | **83*** |
| Jamalpur | 4 | 21 | 44 | 0 | 4 | 11 | 11 | 28 | 48 |
| Jashore | 1 | 5 | 15 | 6 | 22 | 43 | 2 | 7 | 15 |
| Jhalokati | 2 | 7 | 16 | 3 | 11 | 23 | 0 | 0 | 0 |
| Jhenaidah | 0 | 0 | 0 | 0 | 1 | 4 | 0 | 1 | 3 |
| Joypurhat | 0 | 0 | 0 | **73*** | **93*** | **98*** | 28 | **51*** | **69*** |
| Khagrachhari | 0 | 0 | 0 | 0 | 0 | 1 | 0 | 0 | 0 |
| Khulna | 0 | 1 | 5 | 14 | **42*** | **67*** | 1 | 4 | 11 |
| Kishoreganj | 2 | 12 | 27 | 2 | 8 | 22 | 0 | 2 | 6 |
| Kurigram | 1 | 8 | 21 | 2 | 10 | 28 | **37*** | **54*** | **67*** |
| Kushtia | 0 | 0 | 0 | 0 | 1 | 4 | 8 | 21 | 35 |
| Lakshmipur | 0 | 0 | 0 | 0 | 0 | 2 | **88*** | **97*** | **99*** |
| Lalmonirhat | 0 | 0 | 1 | 1 | 7 | 19 | 22 | **46*** | **68*** |
| Madaripur | 0 | 4 | 10 | 1 | 5 | 16 | 9 | 22 | 38 |
| Magura | 3 | 15 | 30 | 0 | 1 | 4 | 0 | 0 | 0 |
| Manikganj | 11 | 30 | **51*** | 0 | 0 | 0 | 0 | 0 | 0 |
| Meherpur | 0 | 0 | 0 | 0 | 0 | 0 | 0 | 0 | 1 |
| Moulvibazar | 7 | 26 | **48*** | 31 | **59*** | **77*** | **62*** | **74*** | **83*** |
| Munshiganj | 9 | 33 | **54*** | 0 | 0 | 0 | 0 | 1 | 4 |
| Mymensingh | 0 | 3 | 9 | 0 | 0 | 3 | 0 | 0 | 0 |
| Naogaon | 29 | **52*** | **67*** | **81*** | **93*** | **97*** | 19 | 37 | **54*** |
| Narail | 0 | 0 | 0 | 1 | 7 | 16 | 1 | 4 | 9 |
| Narayanganj | 0 | 0 | 0 | 0 | 2 | 9 | 0 | 0 | 0 |
| Narsingdi | 0 | 2 | 10 | 0 | 3 | 14 | 0 | 0 | 0 |
| Natore | 1 | 6 | 14 | 18 | **44*** | **67*** | 0 | 1 | 4 |
| Netrokona | **50*** | **82*** | **94*** | 1 | 8 | 19 | 8 | 24 | 46 |
| Nilphamari | 0 | 2 | 8 | 37 | **71*** | **87*** | 18 | **41*** | **62*** |
| Noakhali | **28*** | **69*** | **89*** | 0 | 0 | 3 | 11 | 36 | **64*** |
| Pabna | 4 | 14 | 27 | 0 | 2 | 6 | 1 | 3 | 10 |
| Panchagarh | 0 | 2 | 9 | 1 | 6 | 17 | 15 | 34 | **53*** |
| Patuakhali | 25 | **51*** | **71*** | 13 | 30 | **48*** | 0 | 0 | 1 |
| Pirojpur | 0 | 0 | 0 | 0 | 1 | 6 | **45*** | **69*** | **85*** |
| Rajbari | 0 | 2 | 5 | 0 | 0 | 0 | 0 | 0 | 1 |
| Rajshahi | 0 | 1 | 3 | **83*** | **95*** | **99*** | 0 | 1 | 2 |
| Rangamati | 0 | 2 | 6 | 11 | 28 | **45*** | 3 | 9 | 17 |
| Rangpur | 30 | **59*** | **77*** | 0 | 2 | 8 | 4 | 18 | 38 |
| Satkhira | 0 | 1 | 2 | 0 | 0 | 2 | 23 | **44*** | **63*** |
| Shariatpur | **58*** | **87*** | **96*** | 10 | 29 | **49*** | 0 | 0 | 3 |
| Sherpur | 2 | 18 | 40 | 12 | 36 | **60*** | **75*** | **89*** | **96*** |
| Sirajganj | 1 | 5 | 16 | 5 | 19 | 36 | 6 | 20 | 41 |
| Sunamganj | **99*** | **100*** | **100*** | 0 | 2 | 11 | 0 | 0 | 1 |
| Sylhet | **100*** | **100*** | **100*** | 0 | 4 | 13 | 7 | 23 | 43 |
| Tangail | 1 | 6 | 13 | 7 | 25 | **49*** | **60*** | **78*** | **88*** |
| Thakurgaon | 0 | 0 | 2 | 32 | **62*** | **82*** | 6 | 20 | 42 |
