## Supplementary material for "Geographic prioritization of child nutrition services in Bangladesh: comparing stunting and wasting indicators using the 2025 Multiple Indicator Cluster Survey": S2_Table.docx

**S2 Table. District-level results: hotspots, environmental adjustment and fieldwork month**

**A. Hotspot districts for stunting and wasting**

| Hotspot for | District (division) | Children, n | Prevalence, % (95% CI) | Gi* z | FDR q | Other outcome, % (95% CI) |
| --- | --- | --- | --- | --- | --- | --- |
| Stunting | Sylhet (Sylhet) | 389 | 37.3 (32.1 to 42.6) | 3.71 | 0.007 | Wasting 12.0 (8.2 to 15.7) |
| Stunting | Sunamganj (Sylhet) | 435 | 36.3 (31.1 to 41.6) | 3.71 | 0.007 | Wasting 12.0 (8.6 to 15.4) |
| Stunting | Moulvibazar (Sylhet) | 316 | 26.0 (20.9 to 31.0) | 3.42 | 0.013 | Wasting 16.1 (11.3 to 21.0) |
| Stunting | Netrokona (Mymensingh) | 334 | 30.1 (25.0 to 35.2) | 2.17 | 0.24 | Wasting 12.4 (8.4 to 16.4) |
| Wasting | Naogaon (Rajshahi) | 211 | 20.0 (14.0 to 26.0) | 3.26 | 0.070 | Stunting 27.7 (19.9 to 35.4) |
| Wasting | Chapai Nawabganj (Rajshahi) | 317 | 13.1 (10.0 to 16.1) | 2.85 | 0.14 | Stunting 23.1 (18.9 to 27.3) |

Districts with Gi* z above 1.96 among 64 districts (city corporations included in their parent districts). Prevalence is survey-weighted among children with a valid measurement. FDR q is the Benjamini-Hochberg value across 64 districts. National prevalence was 24.0% for stunting and 12.9% for wasting. Moulvibazar is flagged because neighboring districts have high rates, and its own prevalence is close to the national figure. Habiganj (z=1.95) fell just below the threshold. Chapai Nawabganj is spelled Chapainababganj in the survey files. The three districts passing FDR correction (Sylhet, Sunamganj and Moulvibazar) had a mean 2020 rainfall of 5,358 mm across their clusters, against 2,549 mm in the other 61 districts (MICS 2025 geospatial covariates file).

**B. Generic regional terms compared with the environmental factors**

Stunting

| Adjustment | Terms added | Moran’s I | Permutation P | FDR hotspots | z, Sylhet | Between-district SD | Variance removed (%) |
| --- | --- | --- | --- | --- | --- | --- | --- |
| Observed (no adjustment) | 0 | 0.214 | 0.003 | 3 | 3.71 | 0.034 | 0 |
| Age, sex, wealth, area only | 0 | 0.232 | 0.001 | 3 | 3.60 | 0.037 | −21 |
| Plus six environmental factors | 7 | −0.011 | 0.45 | 0 | 1.13 | 0.021 | 63 |
| Plus rainfall alone | 1 | 0.154 | 0.016 | 0 | 1.69 | 0.031 | 17 |
| Plus five factors without rainfall | 6 | 0.110 | 0.047 | 0 | 2.17 | 0.029 | 25 |
| Plus latitude and longitude quadratic trend | 5 | 0.061 | 0.14 | 0 | 0.36 | 0.031 | 17 |
| Plus division | 7 | −0.084 | 0.83 | 0 | 0.53 | 0.028 | 30 |

Wasting

| Adjustment | Terms added | Moran’s I | Permutation P | FDR hotspots | z, Sylhet | Between-district SD | Variance removed (%) |
| --- | --- | --- | --- | --- | --- | --- | --- |
| Observed (no adjustment) | 0 | 0.052 | 0.17 | 0 | 0.42 | 0.019 | 0 |
| Age, sex, wealth, area only | 0 | −0.006 | 0.43 | 0 | 0.48 | 0.019 | −2 |
| Plus six environmental factors | 7 | −0.063 | 0.73 | 0 | 0.97 | 0.019 | −5 |
| Plus rainfall alone | 1 | −0.009 | 0.44 | 0 | 0.41 | 0.019 | −1 |
| Plus five factors without rainfall | 6 | −0.062 | 0.74 | 0 | 1.06 | 0.019 | −6 |
| Plus latitude and longitude quadratic trend | 5 | −0.043 | 0.63 | 0 | 0.84 | 0.019 | 0 |
| Plus division | 7 | −0.114 | 0.93 | 0 | 0.46 | 0.016 | 26 |

All models were survey-weighted logistic models without division, except in the last row. Each row shows district means of observed minus predicted values, with the same five-nearest-centroid neighbors and 9,999 permutations as the main district analysis. Between-district SD is the square root of the DerSimonian-Laird between-district variance. Variance removed is the in-sample fall in that variance from the observed (unadjusted) value. Measured from the model with age, sex, wealth and area, the six factors removed 69%, division 42%, rainfall alone 32% and the trend 32%. Intervals are in panel C.

**C. Bootstrap intervals for the stunting comparison in panel B**

| Adjustment | Moran’s I (95% CI) | Variance removed, % (95% CI) |
| --- | --- | --- |
| Observed (no adjustment) | 0.214 (0.04 to 0.27) | 0 (0 to 0) |
| Age, sex, wealth, area only | 0.232 (0.06 to 0.29) | −21 (−50 to 7) |
| Plus six environmental factors | −0.011 (−0.12 to 0.09) | 63 (28 to 84) |
| Plus rainfall alone | 0.154 (−0.02 to 0.23) | 17 (−23 to 44) |
| Plus five factors without rainfall | 0.110 (−0.04 to 0.18) | 25 (−5 to 47) |
| Plus latitude and longitude quadratic trend | 0.061 (−0.05 to 0.11) | 17 (−32 to 56) |
| Plus division | −0.084 (−0.15 to −0.01) | 30 (−5 to 63) |

| Comparator | Difference in percentage points (95% CI) | Share of bootstrap samples favouring the six factors |
| --- | --- | --- |
| Rainfall alone | 45 (19 to 77) | 100% |
| Five factors without rainfall | 37 (16 to 57) | 100% |
| Latitude and longitude quadratic trend | 45 (5 to 88) | 99% |
| Division | 33 (−4 to 60) | 96% |

Stratified cluster bootstrap (Rao-Wu rescaling): within each stratum, one fewer primary sampling unit than the stratum contained was drawn with replacement, 2,000 times. Each replicate multiplied the survey weights by the Rao-Wu factor, refitted every model and recomputed district means, their linearized standard errors, the between-district variance and Moran’s I. Negative DerSimonian-Laird estimates were set to zero. Each district mean was weighted by the inverse of its squared standard error. Intervals are 2.5th and 97.5th percentiles. The difference is the percentage of between-district variance removed by the six factors minus that removed by the comparator, in the same replicate. With 64 districts, the intervals are wide. District standard errors treat each model’s predictions as fixed and ignore correlation between districts, while the bootstrap carries the uncertainty from refitting the models. The measure is descriptive.

**D. Weighted prevalence by month of fieldwork**

| Month of measurement | Children (stunting) | Stunting % (95% CI) | Children (wasting) | Wasting % (95% CI) |
| --- | --- | --- | --- | --- |
| Feb | 1,093 | 24.3 (21.2 to 27.3) | 1,090 | 9.7 (8.0 to 11.4) |
| Mar | 6,425 | 25.0 (23.7 to 26.3) | 6,433 | 12.1 (11.1 to 13.1) |
| Apr | 5,804 | 23.4 (22.0 to 24.7) | 5,806 | 13.2 (12.2 to 14.1) |
| May | 6,458 | 23.8 (22.4 to 25.2) | 6,443 | 13.8 (12.8 to 14.7) |
| Jun | 2,540 | 23.4 (21.3 to 25.5) | 2,539 | 13.3 (11.7 to 14.9) |

Month of anthropometric measurement was used where it fell between February and June (23,340 of 24,680 children); otherwise, the child interview date was used. Design-based test of prevalence by month: stunting P=0.50, wasting P=0.007. District mean fieldwork day was correlated with district wasting prevalence (Spearman r=0.37, P=0.003) and not with stunting (r=−0.10, P=0.43). District mean fieldwork day showed spatial dispersion rather than clustering: neighboring districts tended to be surveyed at different times (Moran’s I −0.18, expected −0.016; two-sided permutation P=0.008). Every division was surveyed in all five months. In each division, 4% to 7% of children were measured in February, 21% to 37% in March, 22% to 30% in April, 22% to 36% in May and 7% to 15% in June. Each district’s fieldwork spanned two to five months.
