## Supplementary material for "Geographic prioritization of child nutrition services in Bangladesh: comparing stunting and wasting indicators using the 2025 Multiple Indicator Cluster Survey": S3_Table.docx

**S3 Table. Individual-level models: adjusted and unadjusted associations, altitude form, the stunting and wasting difference, full primary model and goodness of fit**

**A. Adjusted associations, primary model**

| Factor | Outcome | OR | 95% CI | SE | P | FDR-adjusted P | OR (no division) | SE (no division) | P (no division) |
| --- | --- | --- | --- | --- | --- | --- | --- | --- | --- |
| Altitude (per 100 m) | Stunting | 0.64 | 0.44 to 0.92 | 0.189 | 0.017 | 0.177 | 0.55 | 0.143 | <0.001 |
| Altitude (per 100 m) | Wasting | 1.39 | 0.95 to 2.02 | 0.192 | 0.090 | 0.435 | 1.35 | 0.153 | **0.048** |
| Altitude (per 100 m) | Concurrent | 1.18 | 0.62 to 2.27 | 0.332 | 0.612 | 0.835 | NA | NA | NA |
| Motorized travel time (per hour) | Stunting | 0.84 | 0.68 to 1.03 | 0.104 | 0.086 | 0.435 | 0.87 | 0.103 | 0.181 |
| Motorized travel time (per hour) | Wasting | 0.98 | 0.80 to 1.20 | 0.103 | 0.834 | 0.835 | 1.05 | 0.093 | 0.571 |
| Motorized travel time (per hour) | Concurrent | 0.94 | 0.66 to 1.34 | 0.179 | 0.737 | 0.835 | NA | NA | NA |
| PM2.5 (per 10 µg/m³) | Stunting | 0.99 | 0.95 to 1.04 | 0.025 | 0.835 | 0.835 | 1.00 | 0.020 | 0.907 |
| PM2.5 (per 10 µg/m³) | Wasting | 1.02 | 0.96 to 1.09 | 0.031 | 0.475 | 0.832 | 1.02 | 0.025 | 0.439 |
| PM2.5 (per 10 µg/m³) | Concurrent | 1.05 | 0.94 to 1.16 | 0.054 | 0.396 | 0.832 | NA | NA | NA |
| Urbanicity: Town/semi-dense (ref. Rural) | Stunting | 0.90 | 0.77 to 1.06 | 0.080 | 0.213 | 0.638 | 0.85 | 0.077 | **0.035** |
| Urbanicity: Town/semi-dense (ref. Rural) | Wasting | 0.95 | 0.79 to 1.14 | 0.093 | 0.558 | 0.835 | 0.91 | 0.091 | 0.274 |
| Urbanicity: Town/semi-dense (ref. Rural) | Concurrent | 0.93 | 0.67 to 1.28 | 0.163 | 0.644 | 0.835 | NA | NA | NA |
| Urbanicity: City (ref. Rural) | Stunting | 0.94 | 0.78 to 1.12 | 0.092 | 0.475 | 0.832 | 0.93 | 0.089 | 0.441 |
| Urbanicity: City (ref. Rural) | Wasting | 1.02 | 0.83 to 1.26 | 0.106 | 0.832 | 0.835 | 1.02 | 0.105 | 0.840 |
| Urbanicity: City (ref. Rural) | Concurrent | 1.05 | 0.73 to 1.51 | 0.186 | 0.804 | 0.835 | NA | NA | NA |
| Rainfall (per 100 mm) | Stunting | 1.008 | 1.003 to 1.012 | 0.0021 | 0.0005 | **0.010** | 1.013 | 0.0017 | <0.001 |
| Rainfall (per 100 mm) | Wasting | 0.997 | 0.991 to 1.002 | 0.0030 | 0.247 | 0.649 | 1.001 | 0.0023 | 0.609 |
| Rainfall (per 100 mm) | Concurrent | 1.002 | 0.993 to 1.011 | 0.0046 | 0.677 | 0.835 | NA | NA | NA |
| Drought-frequency class (per class) | Stunting | 1.01 | 0.99 to 1.04 | 0.012 | 0.201 | 0.638 | 1.00 | 0.0083 | 0.575 |
| Drought-frequency class (per class) | Wasting | 1.02 | 1.00 to 1.05 | 0.014 | 0.104 | 0.435 | 1.02 | 0.010 | **0.034** |
| Drought-frequency class (per class) | Concurrent | 1.03 | 0.98 to 1.07 | 0.024 | 0.299 | 0.698 | NA | NA | NA |

Estimates are from survey-weighted logistic regression adjusted for child age group, sex, household wealth quintile, area and division. FDR-adjusted P is the Benjamini-Hochberg P value across 21 tests (seven terms, three outcomes). Altitude is shown per 100 m, but its association was not linear (panel C); 95% of clusters lie at or below 55.5 m. The last two columns repeat the model without division, for stunting and wasting only. Bold P (no division) values reached P<0.05 only without division. Bold FDR-adjusted P values are below 0.05. Concurrent estimates are exploratory, given only 902 cases. NA: not run. SE is the survey-linearized standard error of the log odds ratio.

**B. Unadjusted (crude) associations**

| Factor | Outcome | Crude OR | 95% CI | SE | P |
| --- | --- | --- | --- | --- | --- |
| Altitude (per 100 m) | Stunting | 0.88 | 0.69 to 1.12 | 0.123 | 0.304 |
| Altitude (per 100 m) | Wasting | 1.39 | 1.08 to 1.81 | 0.132 | 0.012 |
| Altitude (per 100 m) | Concurrent | 1.67 | 1.14 to 2.44 | 0.193 | 0.008 |
| Motorized travel time (per hour) | Stunting | 1.38 | 1.15 to 1.65 | 0.091 | 0.0004 |
| Motorized travel time (per hour) | Wasting | 1.27 | 1.09 to 1.47 | 0.076 | 0.002 |
| Motorized travel time (per hour) | Concurrent | 1.44 | 1.19 to 1.73 | 0.095 | 0.0001 |
| PM2.5 (per 10 µg/m³) | Stunting | 0.94 | 0.91 to 0.97 | 0.016 | 0.0002 |
| PM2.5 (per 10 µg/m³) | Wasting | 0.98 | 0.94 to 1.01 | 0.019 | 0.207 |
| PM2.5 (per 10 µg/m³) | Concurrent | 0.93 | 0.87 to 0.98 | 0.031 | 0.013 |
| Urbanicity: Town/semi-dense (ref. Rural) | Stunting | 0.78 | 0.67 to 0.92 | 0.080 | 0.002 |
| Urbanicity: Town/semi-dense (ref. Rural) | Wasting | 0.88 | 0.74 to 1.04 | 0.087 | 0.129 |
| Urbanicity: Town/semi-dense (ref. Rural) | Concurrent | 0.80 | 0.59 to 1.07 | 0.150 | 0.130 |
| Urbanicity: City (ref. Rural) | Stunting | 0.70 | 0.59 to 0.82 | 0.084 | <0.0001 |
| Urbanicity: City (ref. Rural) | Wasting | 0.83 | 0.69 to 0.99 | 0.092 | 0.041 |
| Urbanicity: City (ref. Rural) | Concurrent | 0.69 | 0.51 to 0.95 | 0.161 | 0.023 |
| Rainfall (per 100 mm) | Stunting | 1.014 | 1.011 to 1.017 | 0.0015 | <0.0001 |
| Rainfall (per 100 mm) | Wasting | 1.002 | 0.998 to 1.006 | 0.0020 | 0.425 |
| Rainfall (per 100 mm) | Concurrent | 1.009 | 1.003 to 1.014 | 0.0028 | 0.002 |
| Drought-frequency class (per class) | Stunting | 1.01 | 0.99 to 1.02 | 0.0075 | 0.438 |
| Drought-frequency class (per class) | Wasting | 1.02 | 1.00 to 1.04 | 0.0087 | 0.014 |
| Drought-frequency class (per class) | Concurrent | 1.04 | 1.01 to 1.07 | 0.014 | 0.009 |

Estimates are from survey-weighted logistic regression with one factor at a time and no other adjustment. These are shown alongside the adjusted estimates in panel A, as STROBE recommends. SE is the survey-linearized standard error of the log odds ratio.

**C. Altitude associations by functional form**

| Analysis | Stunting OR (95% CI) | SE | P | Wasting OR (95% CI) | SE | P |
| --- | --- | --- | --- | --- | --- | --- |
| **Primary model, six factors** |  |  |  |  |  |  |
| Linear, per 100 m (main analysis) | 0.64 (0.44 to 0.92) | 0.189 | 0.017 | 1.39 (0.95 to 2.02) | 0.192 | 0.090 |
| Per doubling of altitude | 0.96 (0.91 to 1.02) | 0.029 | 0.18 | 1.07 (1.00 to 1.15) | 0.034 | 0.037 |
| Children above 89.3 m excluded (top 1% of children), per 100 m | 0.56 (0.36 to 0.85) | 0.218 | 0.007 | 1.76 (1.06 to 2.90) | 0.255 | 0.027 |
| Clusters above 55.5 m excluded (top 5% of clusters), per 100 m | 0.58 (0.32 to 1.08) | 0.313 | 0.085 | 3.63 (1.83 to 7.22) | 0.350 | 0.0002 |
| Test of non-linearity (quadratic and cubic terms) |  |  | 0.0011 |  |  | 0.0044 |
| **Stacked model: ratio of odds ratios, wasting relative to stunting** | **Ratio (95% CI)** | **SE** | **P** |  |  |  |
| Linear, per 100 m (main analysis) | 2.00 (1.40 to 2.86) | 0.183 | 0.00015 |  |  |  |
| Per doubling of altitude | 1.10 (1.04 to 1.16) | 0.027 | 0.0005 |  |  |  |
| Clusters above 55.5 m excluded, per 100 m | 3.14 (1.83 to 5.39) | 0.275 | 0.00003 |  |  |  |
| Natural spline, 3 degrees of freedom (joint test of outcome-by-altitude terms) |  |  | 0.0006 |  |  |  |

Estimates are from survey-weighted logistic regression adjusted for child age group, sex, household wealth quintile, area and division. The primary model includes all six factors. The stacked rows use the simpler stacked model, in which only altitude and rainfall differ by outcome. Cluster altitude was highly skewed (median 11.0 m, 95th percentile 55.5 m, maximum 233.2 m), so only 5% of clusters lie above 55.5 m and odds ratios per 100 m describe few clusters. Sample sizes after excluding the top 1% were 22,092 (stunting) and 22,081 (wasting); after excluding the top 5% they were 21,137 and 21,130. SE is the survey-linearized standard error of the log odds ratio, or of the log ratio of odds ratios in the stacked rows.

**D. Robustness of the difference between stunting and wasting in their associations with altitude and rainfall**

| Analysis | Altitude: ratio of odds ratios (95% CI) | SE | P | Rainfall: ratio of odds ratios (95% CI) | SE | P |
| --- | --- | --- | --- | --- | --- | --- |
| Fully interacted model, main analysis (all six factors and all adjusters differ by outcome) | 2.18 (1.30 to 3.65) | 0.264 | 0.0033 | 0.989 (0.982 to 0.996) | 0.0036 | 0.0021 |
| Without division adjustment | 2.04 (1.42 to 2.93) | 0.185 | 0.00012 | 0.986 (0.981 to 0.991) | 0.0026 | <0.001 |
| Standard errors clustered on 67 survey domains | 2.00 (1.32 to 3.04) | 0.207 | 0.0017 | 0.986 (0.980 to 0.992) | 0.0031 | <0.001 |
| Simpler stacked model (only altitude and rainfall differ by outcome) | 2.00 (1.40 to 2.86) | 0.183 | 0.00015 | 0.986 (0.981 to 0.991) | 0.0026 | <0.001 |

Ratios compare wasting with stunting. The rows without division and with domain-clustered errors use the simpler stacked model. In the fully interacted model the other factors did not differ by outcome: motorized travel time 1.17 (0.88 to 1.56), P=0.29; PM2.5 1.03 (0.95 to 1.11), P=0.48; town or semi-dense urbanicity 1.05 (0.82 to 1.33), P=0.71; city 1.09 (0.83 to 1.43), P=0.52; drought 1.01 (0.97 to 1.04), P=0.68. The joint test of all seven factor-by-outcome terms gave P=0.0039. The Bonferroni threshold for seven tests is 0.007. SE is the survey-linearized standard error of the log ratio of odds ratios.

**E. Full primary model: all coefficients**

| Term | Stunting OR (95% CI) | SE | P | Wasting OR (95% CI) | SE | P | Concurrent OR (95% CI) | SE | P |
| --- | --- | --- | --- | --- | --- | --- | --- | --- | --- |
| Intercept (odds for the reference child) | 0.29 (0.17 to 0.50) | 0.279 | <0.001 | 0.15 (0.07 to 0.29) | 0.350 | <0.001 | 0.006 (0.002 to 0.023) | 0.655 | <0.001 |
| **Environmental and access factors** |  |  |  |  |  |  |  |  |  |
| Altitude (per 100 m) | 0.64 (0.44 to 0.92) | 0.189 | 0.017 | 1.39 (0.95 to 2.02) | 0.192 | 0.090 | 1.18 (0.62 to 2.27) | 0.332 | 0.612 |
| Motorized travel time (per hour) | 0.84 (0.68 to 1.03) | 0.104 | 0.086 | 0.98 (0.80 to 1.20) | 0.103 | 0.834 | 0.94 (0.66 to 1.34) | 0.179 | 0.737 |
| PM2.5 (per 10 µg/m³) | 0.99 (0.95 to 1.04) | 0.025 | 0.835 | 1.02 (0.96 to 1.09) | 0.031 | 0.475 | 1.05 (0.94 to 1.16) | 0.054 | 0.396 |
| Urbanicity: Town/semi-dense (ref. Rural) | 0.90 (0.77 to 1.06) | 0.080 | 0.213 | 0.95 (0.79 to 1.14) | 0.093 | 0.558 | 0.93 (0.67 to 1.28) | 0.163 | 0.644 |
| Urbanicity: City (ref. Rural) | 0.94 (0.78 to 1.12) | 0.092 | 0.475 | 1.02 (0.83 to 1.26) | 0.106 | 0.832 | 1.05 (0.73 to 1.51) | 0.186 | 0.804 |
| Rainfall (per 100 mm) | 1.008 (1.003 to 1.012) | 0.0021 | <0.001 | 0.997 (0.991 to 1.002) | 0.0030 | 0.247 | 1.002 (0.993 to 1.011) | 0.0046 | 0.677 |
| Drought-frequency class (per class) | 1.01 (0.99 to 1.04) | 0.012 | 0.201 | 1.02 (1.00 to 1.05) | 0.014 | 0.104 | 1.03 (0.98 to 1.07) | 0.024 | 0.299 |
| **Child age group (ref. 0 to 5 months)** |  |  |  |  |  |  |  |  |  |
| 6 to 11 months | 0.99 (0.82 to 1.19) | 0.097 | 0.898 | 0.91 (0.73 to 1.12) | 0.108 | 0.361 | 2.41 (1.35 to 4.29) | 0.295 | 0.003 |
| 12 to 23 months | 2.01 (1.72 to 2.35) | 0.080 | <0.001 | 0.94 (0.78 to 1.12) | 0.091 | 0.475 | 6.06 (3.68 to 9.97) | 0.254 | <0.001 |
| 24 to 35 months | 2.03 (1.73 to 2.38) | 0.082 | <0.001 | 0.96 (0.81 to 1.15) | 0.091 | 0.686 | 5.23 (3.17 to 8.64) | 0.256 | <0.001 |
| 36 to 47 months | 1.71 (1.45 to 2.01) | 0.082 | <0.001 | 1.00 (0.83 to 1.20) | 0.095 | 0.974 | 5.72 (3.49 to 9.38) | 0.252 | <0.001 |
| 48 to 59 months | 1.34 (1.14 to 1.58) | 0.083 | <0.001 | 1.09 (0.91 to 1.30) | 0.091 | 0.370 | 5.12 (3.10 to 8.46) | 0.256 | <0.001 |
| **Sex (ref. male)** |  |  |  |  |  |  |  |  |  |
| Female | 0.92 (0.85 to 0.99) | 0.038 | 0.029 | 0.82 (0.75 to 0.90) | 0.046 | <0.001 | 0.80 (0.68 to 0.94) | 0.082 | 0.006 |
| **Household wealth quintile (ref. poorest)** |  |  |  |  |  |  |  |  |  |
| Second | 0.75 (0.67 to 0.83) | 0.056 | <0.001 | 1.04 (0.91 to 1.20) | 0.071 | 0.579 | 0.83 (0.68 to 1.02) | 0.104 | 0.075 |
| Middle | 0.64 (0.57 to 0.71) | 0.058 | <0.001 | 0.85 (0.73 to 0.98) | 0.076 | 0.028 | 0.68 (0.53 to 0.86) | 0.124 | 0.002 |
| Fourth | 0.55 (0.49 to 0.61) | 0.059 | <0.001 | 0.87 (0.75 to 1.01) | 0.076 | 0.058 | 0.54 (0.42 to 0.70) | 0.130 | <0.001 |
| Richest | 0.36 (0.32 to 0.42) | 0.070 | <0.001 | 0.62 (0.52 to 0.74) | 0.086 | <0.001 | 0.40 (0.29 to 0.54) | 0.155 | <0.001 |
| **Area (ref. urban)** |  |  |  |  |  |  |  |  |  |
| Rural | 0.99 (0.89 to 1.09) | 0.052 | 0.821 | 1.00 (0.89 to 1.11) | 0.057 | 0.951 | 0.99 (0.81 to 1.20) | 0.099 | 0.910 |
| **Division (ref. Barishal)** |  |  |  |  |  |  |  |  |  |
| Chattogram | 1.20 (1.02 to 1.41) | 0.083 | 0.027 | 1.14 (0.93 to 1.39) | 0.101 | 0.209 | 1.46 (0.99 to 2.16) | 0.200 | 0.058 |
| Dhaka | 1.06 (0.90 to 1.23) | 0.080 | 0.500 | 0.92 (0.75 to 1.12) | 0.102 | 0.409 | 1.04 (0.71 to 1.52) | 0.195 | 0.857 |
| Khulna | 0.81 (0.67 to 0.98) | 0.097 | 0.031 | 0.91 (0.72 to 1.15) | 0.120 | 0.439 | 1.23 (0.81 to 1.87) | 0.214 | 0.333 |
| Mymensingh | 0.93 (0.76 to 1.14) | 0.105 | 0.487 | 0.90 (0.69 to 1.18) | 0.137 | 0.464 | 1.14 (0.73 to 1.79) | 0.229 | 0.564 |
| Rajshahi | 0.95 (0.77 to 1.16) | 0.104 | 0.587 | 1.06 (0.83 to 1.35) | 0.122 | 0.632 | 1.52 (0.98 to 2.34) | 0.221 | 0.059 |
| Rangpur | 0.96 (0.77 to 1.20) | 0.112 | 0.745 | 1.00 (0.77 to 1.29) | 0.130 | 0.975 | 1.37 (0.88 to 2.14) | 0.228 | 0.169 |
| Sylhet | 1.31 (1.05 to 1.64) | 0.113 | 0.016 | 1.26 (0.94 to 1.70) | 0.151 | 0.122 | 1.95 (1.17 to 3.26) | 0.262 | 0.011 |

This panel gives every coefficient of the primary model in panel A for the three outcomes. The model is a survey-weighted logistic regression of the six factors with child age group, sex, household wealth quintile, area and division. The factor rows repeat panel A. The intercept is the odds of the outcome for a child in every reference category with all continuous factors at zero. SE is the survey-linearized standard error of the log odds ratio, or of the log odds for the intercept. Concurrent estimates are exploratory, given only 902 cases.

**F. Goodness of fit of the primary models**

| Measure | Stunting | Wasting | Concurrent |
| --- | --- | --- | --- |
| Children analysed | 22,320 | 22,311 | 22,205 |
| Cases | 5,365 | 2,905 | 902 |
| Primary sampling units | 3,144 | 3,144 | 3,144 |
| Estimated coefficients | 26 | 26 | 26 |
| Archer-Lemeshow F (df) | 0.76 (9, 3,004) | 0.72 (9, 3,004) | 0.37 (9, 3,004) |
| Archer-Lemeshow P | 0.658 | 0.693 | 0.952 |
| Nagelkerke pseudo-R² | 0.050 | 0.011 | 0.038 |
| Cox-Snell pseudo-R² | 0.034 | 0.006 | 0.011 |
| Design-based AIC | 23,913.6 | 17,074.5 | 7,281.9 |

Measures are for the primary models in panels A and E. The Archer-Lemeshow test regresses each child's residual (outcome minus predicted probability) on ten survey-weighted deciles of predicted probability. It uses a design-based Wald F test, and a large P value gives no evidence of poor calibration. Pseudo-R² values use the survey weights. The design-based Akaike information criterion (AIC) can be compared only between models fitted to the same children.
